# A rapid review of what organisational level factors support or inhibit the scale and spread of innovations in children’s social care

**DOI:** 10.1101/2023.04.03.23288061

**Authors:** Mala Mann, Kate Lifford, Susan O’Connell, Alison Weightman, Lydia Searchfield, Ruth Lewis, Alison Cooper, Adrian Edwards

## Abstract

Innovation may provide a means for tackling challenges facing childrens social care, some of them deep-rooted and many exacerbated by COVID-19. Welsh Government has recently committed to a significant 3-year investment to support innovation in adults and childrens social care. The delivery of social care in Wales has a complex and multi-faceted approach, involving collaborative working between a range of organisations, which will likely affect decisions around implementation and scale-up of new and/or existing interventions. The aim of the review was to identify any factors (barriers and enablers) that affect the implementation and scale up of an innovation in childrens social care organisations.

Ten studies were identified, comprising three secondary studies (reviews) and seven primary studies. Factors potentially influencing scale and spread of innovation were extracted and categorised. The domains (and sub-domains) covered by included studies were; adopters (staff role/identity; carer input), organisation (capacity to innovate; readiness for change; nature of adoption/funding; extent of change needed; work needed to implement), and wider system (political/policy; regulatory/legal; professional; socio-cultural).

Enablers for which a clear consensus seems to be emerging across the literature included: specific training and support for professional staff, support and mutual respect within inter-professional and professional-carer relationships, senior management/leadership buy-in and support, multi-disciplinary communication and joint working, and developing compatible data systems to support joint working/collaboration. Barriers for which a clear consensus seems to be emerging across the literature were: short term or lack of funding (the need for funding was presented as an enabler in some studies), and implementation difficulties (e.g. multiple priorities and changing structures).

Policy Implications: This review highlights the complexity of the social care models but provides some clear pointers for policy and practice. The findings indicate the need for: senior management buy-in and support, short and longer term funding, multi-disciplinary communication and joint working, good professional (and professional-carer) relationships with support and mutual respect, and specific training and support for professional staff.

The confidence in the evidence is uncertain as the study designs included non-systematic reviews and service evaluations; most studies did not use a formal methodology and all had some quality limitations.

## 1. BACKGROUND

### 1.1 Who is this review for?

This Rapid Review is being conducted as part of the Health and Care Research Wales Evidence Centre Work Programme. The review was requested by Social Care Wales (SCW) Children’s Social Care Sector and will be useful for informing the work at SCW and for the children’s social care sector.

### 1.2 Background and purpose of this review

#### 1.2.1 Purpose of review

Social Care Wales (SCW) have a major programme of work on supporting innovation and as part of that will be engaging with the children’s social care sector on their priorities. This rapid review will provide insight into what needs to be in place for innovation to be adopted, spread, and scaled. SCW plan to use the findings to inform the work of their new team members (Community Managers, Innovation Coaching Manager, Innovation Coaches, and Senior Evaluation Lead), who will be working directly with practitioners to support innovation. The rapid review will highlight what targeted support might be needed, and the key issues they should bring people together on. The SASCI (Supporting Adult Social Care Innovation) project led by London School of Economics has provided similar insights for adult social care, but this work will ensure SCW understand what is needed from a children’s social care perspective.

The timing of this rapid review is particularly pertinent, because children’s services are having to consider new ways of working in light of the First Minister’s pledge to eliminate profit in children’s social care. Given that much of the children’s residential care market in Wales is privately operated, this will require a step change in the way services are run, with innovation at the heart of this change. SCW anticipate that the findings from this rapid review will be useful to the sector in navigating these changes and will use their channels to share the findings widely.

Factors potentially influencing scale and spread of innovation were categorised into three domains for this review: adopters, organisation and wider system (see Section 5.4). The domains of adopters and wider system were included along with organisation because of their relevance. Adopters included professionals and professional-carer relationships within organisations, Wider system covered inter-organisation factors.

#### 1.2.1 Social care in Wales

Social care is a devolved issue and as such there are variations across the UK. In Wales, the Welsh Parliament is responsible for legislating for children’s social care. Delivering social care in Wales has a complex and multi-faceted approach, involving collaborative working between a range of organisations. Describing the social care delivery model for children in Wales is not within the scope of this rapid review. However, it is important to understand that, while there is a legal framework which covers the provision of children’s social care services in the form of the Social Services and Well-being (Wales) Act 2014, there are variations in how services are implemented and provided across Wales which may affect decisions around implementation and scale-up of new and/or existing interventions.

Web searching and personal contact with local authorities (LAs) within Wales have highlighted the complexity of social care delivery including:

- A high degree of collaboration between LAs, local health boards and third sector (charity) organisations
- Unclear structure for delivery – who delivers what and how?
- Unclear funding structures
- Some initiatives identified as ongoing in Wales had little or no information available (See Section 2.2 ‘The Wales Perspective’)
- Similar interventions and/or services being implemented in different regions but being called something different (for the No Wrong Door approach discussed below).
- A lack of available evaluations

While initiatives and programmes funded by the third sector are not included in the scope of this review, many of the initiatives identified are in some way aligned with third sector partners which makes it difficult to exclude them completely.

Through the Social Services and Well-being (Wales) Act 2014, seven statutory regional partnership boards (RPBs) were set up in 2016 with the aim of driving strategic regional delivery of social services in close collaboration with health. RPBs bring together LAs, local health boards and third sector to address the health and social care needs of their populations. The RPBs are required by law to prioritise the integration of services for children with complex needs with a focus on preventative services for children and families and care and support services for children that require it – the aim being to prevent the child becoming looked after or enter custody.

RPBs decide their overarching approach to children’s social care for their region but there appear to be some specific requirements set out that they must include. Guidance and support in the form of planning tools, examples of best practice and resources are available for RPBs to help them to deliver their service. An example of this is the NEST Framework (https://nestwales.org), a planning tool that aims to ensure a ‘whole system’ approach for children’s social care and includes an approach called ‘No Wrong Door’ as one of its key principles. The ‘No Wrong Door’ approach is where professionals that offer extra support come together to work out what and who can help most. In 2019/20 the Children’s commissioner for Wales reported that every RPB had a plan for children’s provision and had begun making changes towards a No Wrong Door Approach. A Welsh implementation of ‘No Wrong Door’ (Gwent SPACE-Wellbeing) is included as one of the Welsh innovations in Section 2.2.

## 2. RESULTS

### 2.1 Overview of the Evidence Base

Of the 195 records that were screened at full text, ten studies (reported in 13 publications) from secondary and primary research were included in this review, comprising three secondary studies (reviews) and seven primary studies that were not included in the reviews (see Flow diagram in Section 6).

There are three secondary studies of relevance to the UK: one systematic review (Sheehan et al. 2018); one non-systematic overview of four innovations (Godar & Botcherby 2020); one systematic assessment of the Department for Education funded Children’s Social Care Innovation Programmme (CSCIP) projects Phase 1 (Sebba et al. 2017a), Phase 2 (FitzSimons et al. 2020) and related practice review of Partners in Practice (PIP; Ruch & Maglajlic 2020) (Section 2.1.1). In the protocol for this review there was an intention to exclude secondary research studies, but these three studies were deemed relevant since they provide a rich body of evidence, from a very large number of primary studies, of direct relevance to the scale and spread of innovations in the UK.

There were seven primary studies not included in the reviews described above.

Five of the primary research studies were carried out in a UK setting outside Wales. Three with a qualitative study design (Alderson et al. 2022, Oliveira et al. 2022, Turney & Ruch 2018); one multi-component evaluation (Ecorys UK, 2017); and one mixed methods service evaluation (Plumridge & Sebba 2018). (Section 2.1.2).

Two of the primary research studies were carried out in Wales; one qualitative study (Rees & Handley 2022) and one service evaluation (Shelton et al. 2020). For this review we also included five additional innovations implemented in Wales to enhance the Wales perspective. Two with unpublished evidence of factors that may be related to scale and spread, and a further three where the only evidence relating to scaling of the innovation, to date, comes from elsewhere in the UK (Section 2.1.3).

#### 2.1.1 Secondary evidence

The three reviews are summarised below and with additional study detail and quality assessment in presented in Section 6.2.

Godar & Botcherby (2020) provides a non-systematic overview of four innovation projects (**Achieving Change Together; No Wrong Door; Stockport Family and Team Around the School; Salford Strengthening Families**) that were being used in ten LAs and were part of the Greater Manchester **Scale and Spread programme**. An iterative, collaborative approach was taken to selecting innovations to adopt. The innovation leads worked with LAs to support scale-up by providing coaching and resources. Qualitative data was collected to explore barriers and enablers. Organisational factors enabling implementation of the innovations included **support and attention from senior staff** and **funding** (start up and sustainability). Provision of opportunities for staff to reflect, research, and develop was an enabler at the level of adopters.

Study quality: Not applicable.

Sebba et al. (2017a), FitzSimons et al. (2020), Ruch & Maglajlic (2020)

We identified two related summary reports published by the Department for Education, evaluating Phases 1 and 2 of the **Children’s Social Care Innovation programme (CSCIP)** (Sebba et al. 2017a, FitzSimons et al. 2020). A third related summary report provided more detail on the **Partners in Practice (PIP) programme** which ran alongside the CSCIP (Ruch & Maglajlic 2020). The CSCIP funded a number of innovation projects in England for supporting children who need help from children’s social care services. More information about the CSCIP and PIP along with individual evaluation reports can be found here: https://www.gov.uk/guidance/childrens-social-care-innovation-programme-insights-and-evaluation. It should be noted that some of the examples from the Welsh perspective include innovations which were first implemented in England as part of the CSCIP (See Section 2.2).

Sebba et al. (2017a) summarises the findings from the individual evaluations of **56 innovation projects** across England, identifying a range of ‘hard’ and ‘soft’ outcomes for comparison across the different interventions. Key enablers and barriers to the adoption, scale and spread of these innovations were reported (further details are given in a thematic report^1^). FitzSimons et al. (2020) summarises the findings of independent evaluations of **47 projects** from **Phase 2** of the CSCIP, as well as those from **eight ‘light-touch’** follow up evaluations of Phase 1 projects and a further **seven Phase 1 projects** which became **PIPs**. They report that many of the findings were congruent with those from Phase 1 and noted that **knowledge gained about barriers and enablers had not always been used effectively** by the Phase 2 projects. Challenges for the evaluations included limited evaluation periods, small cohorts, lack of comparator data and data quality concerns. Ruch & Maglajlic (2020) provided a further summary of the experiences of PIPs from both Round 1 and Round 2 of the PIP programme. The PIP programme (commencing 2016) ran alongside the CSCIP. The aim of the PIP programme was to create a partnership between local and central government to improve the children’s social care system by bringing together practitioners and leaders from areas with excellent practice. While seven LA Children’s Services were designated PIP status in Round 1 of the PIP programme and nine additional in Round 2 for a total of 16 with data from 14 PIPs included in the report.

Across these three studies (i.e., noted within two or more studies) a number of factors relevant to organisational support were identified, including the importance of **strong leadership**, **adequate and sustained resourcing**, **multi-agency collaboration** and joint working, and being **realistic in planning** and setting modest goals. **Organisational resistance to change,** was also identified across the studies as a barrier, with services noting that system-level changes happened slowly. Other organisational factors included **systems to support data sharing** (enabler) and **balancing different aspects** of the innovation work (barrier). Knowledge and skills training for practitioners was identified across the studies as an enabler at the level of adopters. Considering embedding researchers and having good professional relationships were also enablers at the adopter level. Supporting changes in whole system professional practice was identified across the studies as a wider system factor. Deregulation in the wider system was identified as an important ongoing policy change which supported innovation.

Study quality: Unclear, no formal critical appraisal tool applicable to study type.

Sheehan et al. (2018) completed a systematic review and realist synthesis of **Signs of Safety** (SoS), a strengths-based, safety-organised approach to collaborative child protection case work. Thirty-eight publications were included (13 from the UK, including one from Wales) and the remaining from other countries that the authors deemed to be relevant to the UK (e.g., USA, Canada). The majority of included studies were qualitative (all UK studies were qualitative), but there were five intervention studies. At an organisational level, factors which influenced implementation included it being **‘organisation led’** with **active leadership**, **multi-organisation culture change,** and **data recording systems to support sharing practice.** Modelling strengths-based practice by managers also influenced implementation.

Study quality: Low.

#### 2.1.2 Primary evidence

Five primary studies were identified in England and Scotland with potential implications for the spread and scale-up of innovations. Only one of the primary studies (Oliveira 2022) was an adaptation of an innovation used elsewhere, the rest were of the initial implementation of innovations. The five studies are summarized below and with additional study detail and quality assessment in Section 6.2.

Alderson et al. (2022) conducted a qualitative study in Newcastle, England, to examine implementation, service delivery and perceived impact of the **Innovation Pilot Project.** The project aimed to reduce fragmentation between services and increase the identification of children affected by parental alcohol misuse by using a whole family approach. As part of the project, child welfare services were brought together to improve collaboration and communication. Interviews were conducted with family members and staff, along with staff focus groups. Organisational enablers included allowing **time for the team to ‘bed in’** and **establishing a multi-agency recording system**. Having clear staff roles and responsibilities was an enabler to implementation at the level of adopters.

Study quality: Medium.

Ecorys UK (2017) led a group evaluating the **Dundee Early Intervention Team’s Improving Futures project**. This was part of the wider Improving Futures project which was carried out across the UK to test different Voluntary and Community Sector led approaches. The Dundee Early Intervention Team’s Improving Futures project aimed to establish a support service for those who did not meet the threshold for statutory intervention before they reached crisis point. The focus of the service was early intervention and prevention, with a view to complement what Dundee Children’s Services offer. The evaluation had multiple components including project documentation and monitoring data, and qualitative interviews. **Short-term funding was both an organisational barrier and enabler**. It was a barrier by diverting staff time and referral changes which resulted in fewer families receiving support. However, it was an enabler because the service was more effective by prioritising help for families most in need of support. This revised referral process was therefore an important adaptation over time. **Partnership working** was another organisational enabler as was flexible staff working (so they could provide support to families at key times of the day). Good working relationships with wider services to ensure the project fit with other services was a wider system factor. Intervention specific staff training was an important enabler at the level of adopters. Relevant to embedding and adaptation over time is the plan for the service to link with a similar one for younger children.

Study quality: Unclear, no formal critical appraisal tool applicable to study type.

Oliveira et al. (2022) undertook qualitative interviews in a scoping study as part of a wider feasibility and pilot RCT of the **Video-feedback Intervention to promote Positive Parenting and Sensitive Discipline, Foster Care (VIPP-FC)** in England. In the wider study the intervention was first adapted from the original VIPP and the version already adapted for foster children in the Netherlands and tested using a case series. The original intervention is an effective treatment approach for attachment problems in looked-after children. It was being adapted as a treatment approach for foster children who present with reactive attachment disorder symptoms. Because the intervention was being tested with a view to conducting an RCT, factors relate to influencing implementation within a research study (as opposed to within a service), some caution is therefore warranted with the findings because some are about the interface between research and services. For example, the study was not mandatory, thus could be put on the ‘backburner’. Findings highlighted here are those that are more general or seem to transfer across into a practice/service setting. At an organisational level, barriers included a **lack of resources** (predominantly in the LAs) for research, **change and organisational inertia**, and **difficulties in the professional network** in implementing the study. Suggested enablers included dedicating resources to maintain relationships **good face-to-face communication** across the study (including support at every level) and **sharing information/learning** between the research team, LA and wider network. The **framing of the intervention** was also suggested to be important. Lack of standardised organisational structures were also noted as wider system barriers to research. Investing in relationships and the training being viewed as opportunities for staff development were both enablers at the level of adopters. However, lack of understanding about the study (recruitment process, aims, approach, benefits) and the instability of placements were both barriers to recruitment. Being clear about the potential benefits to foster carers was an enabler, whereas their available time was a challenge.

Study quality: Medium.

Plumridge & Sebba (2018) conducted an evaluation of Birmingham City Council’s **Step Down Programme** to identify whether and how the project supports young people and also to consider what works well/less well and what outcomes are achieved for young people. The Step Down Programme aimed to move young people from residential homes into foster (stable) placements. A social investor pays for the additional services upfront, which they receive back for the young people who have stable placements (i.e., in placement for 52 weeks). Quantitative service delivery data was collected as well as qualitative interviews. **Consistent involvement of LA staff** was an organisational enabler, as was **proactive team use of progress meetings** (part of the programme). At the level of adopters, the importance of particular staff roles was highlighted, as well as carers being adequately equipped and supported.

Study quality: Unclear, no formal critical appraisal tool applicable to study type.

Turney & Ruch (2018) explored the contribution of the **Cognitive and Affective Supervisory Approach** (CASA) to social works practitioners’ assessments and decision-making practice in two LAs in England. CASA is a practice-based approach aiming to enhance the quantity and quality of available decision-making information and uses cognitive interviewing techniques originally designed to gain evidence from crime witnesses and victims. The study used qualitative methods including interviews and discussion groups. At an organisational level, **introducing the approach was reported as a difficult task/process**, though benefits were noted.

Study quality: Low.

### 2.2 The Wales perspective

Seven interventions implemented in Wales have been identified that provide some evidence regarding organisational guidance for scaling innovations in children’s social care, either from Welsh evaluation or elsewhere in the UK.

Two have relevant publications with local evaluation data:

1. Rees et al. (2019) Fostering Wellbeing
2. Shelton et al. (2020) Adopting Together Service (initial findings)

Two have some informal evidence of factors that may be related to scaling innovations in Wales, based on web searching and personal contacts:

3. Family Drug and Alcohol Courts
4. Gwent SPACE-Wellbeing

A further three have been implemented in Wales but the only evidence relating to scaling of the innovation, to date, comes from elsewhere in the UK:

5. Family Group Conferencing
6. Mockingbird Family Model
7. Signs of Safety

Each of these seven interventions is summarised below with additional study detail and quality assessment in Table 5 (Section 6.2).

Other initiatives were identified as ongoing in Wales, but no additional information could be identified. These include the Getting Ready Project (Voices from Care Cymru), Creating Space for Change (Pause intervention), Friends 4 U (Cardiff Council), KEEP (North East Wales), Shared Lives Care Leavers (eight sites in Wales).

#### Innovations in Wales with formal local evaluation

##### 1: Fostering Wellbeing

Delivered by the Fostering Network (https://www.thefosteringnetwork.org.uk), **Fostering Wellbeing** is a multi-agency programme funded by Welsh Government. It is based on the successful aspects of two pilot projects, Fostering Network’s Head, Heart, Hands and the London Fostering Achievement Programme to create a hybrid model encompassing three strands of work:

- A set of five themed masterclasses delivered to multi-disciplinary members working in the team around the child (social work, health, education, youth justice).
- The development of the Pioneer foster carer role to provide training, operate a telephone helpline and run support groups for foster carers, based within the LA fostering team offices.
- Service support and action plan.

It was set in a Welsh context and piloted in Cwm Taf from 2017-2019 after which it was rolled out across Wales with continued evaluation of the pan-Wales programme (Rees & Handley 2022).

*Factors of relevance to innovation roll out across institutions in Wales:*

i. **Professional relationships and respect** – For this particular intervention, the importance of valuing foster carers as equal team members.
ii. **Communication and multi-disciplinary learning** – For this intervention, having all members of the team around the child attending masterclasses together.

Study quality: Low.

##### 2: Adopting Together Service

Shelton et al. (2020) report on a preliminary evaluation of the **Adopting Together Service,** which uses a partnership approach to public procurement of childcare service delivery. The service is a collaboration between three voluntary adoption agencies (St David’s Children Society, Barnardo’s Cymru and Adoption UK) and the statutory sector across Wales. Led by St David’s Children Society, it aims to have early intervention and prevention at its core, enabling lifelong placements for children. It was set up in 2016 in response to the growing gap between the number of adopters and number of children awaiting adoption. There were two components to creating the service: developing an effective service that minimises the likelihood of family breakdown and engaging the two sectors (voluntary and statutory) though innovative collaborative practices. To achieve this, the following were needed: 1) implementation of an enhanced support service; 2) creation of working structures for partnership working; 3) maximising knowledge exchange.

*Factors of relevance to innovation roll out across institutions in Wales:*

i. Taking a flexible and collaborative approach with buy-in/engagement.
ii. SWOT^2^ and PESTLE^3^ analyses by organisation to create appropriate structures and separate core business from service work.
iii. Developing a joint relationship management plan
iv. Creating **pragmatic service level agreements** between agencies (voluntary adoption agencies and statutory sector in this case).
v. **Consultation** with stakeholders.

Study quality: Unclear, no formal critical appraisal tool applicable to study type.

#### Innovations in Wales with informal local evaluation

##### 3: Family Drug and Alcohol Courts (FDAC)

FDAC is an evidence-based intervention (Zhang et al. 2019) designed to help parents involved in court-based care proceedings to overcome the substance misuse that has put their children at risk of serious harm. These courts are currently being evaluated by CASCADE (Nov 2021 to Jan 2024) within the South East Wales Local Family Justice Board: https://cascadewales.org/research/evaluation-of-the-family-drug-and-alcohol-court-in-wales-pilot/.

Despite the lack of a formal evaluation to date, the implementation of FDACs illustrates the role of government evaluation and recommendation in driving the adoption of innovation in a Welsh context. The timeline was as follows.

1. Evidence of intervention effectiveness published (Zhang et al. 2019).
2. Commission on Justice in Wales recommended roll out to Wales in Oct 2019 (recommendation 35): https://www.gov.wales/commission-justice-wales-report
3. Business case for roll out to England and Wales published by the Centre for Justice Innovation in September 2021: https://justiceinnovation.org/publications/rolling-out-family-drug-and-alcohol-courts-fdac-business-case
4. Adopted by Welsh Government (Cabinet Paper Nov 2021): https://gov.wales/family-drug-and-alcohol-courts-html. Pilot awarded to South East Wales Local Family Justice Board for period to July 2023 with evaluation by CASCADE, Cardiff University. Contact: David Westlake. The Welsh Government noted that *’If the pilot is deemed successful then lessons from this can be used to support the extension of the FDAC model to other areas in Wales. The evaluation will explore if the FDAC pilot is implemented as intended and whether it operates in a way that enables it to be easily scaled’*.

*Factors of relevance to innovation roll out across institutions in Wales:*

**Regulatory/policy changes** (within the wider system) can drive forward innovation within the sector.

##### 4: Gwent SPACE-Wellbeing (No Wrong Door)

While there appears to be a level of consistency across the Welsh RPBs in terms of the broad approaches to social care, for example the requirement to implement a No Wrong Door Approach, this seems to be coupled with a level of flexibility to develop and implement a model of care which best meets the needs of their specific region. Examples of this include Gwent SPACE-Wellbeing and Start Well in Powys.

As part of its Iceberg Programme, Gwent RPB has developed a model of Single Point of Access for Children’s Emotional Wellbeing and Mental Health panels called SPACE-Wellbeing and Early Help panels. The panels are in place across the five LA areas of Gwent. The panels meet once a week and take referrals from GPs, schools, social services, parents and families. Attendees at the panel include representatives from a wide range of services. The model was evaluated for the Welsh Government as part of a wider evaluation of the Iceberg Transformation Model and is awaiting publication on the web site (Children’s Commissioner for Wales 2023).

Personal communication from members of the Gwent SPACE Wellbeing and Early Help Panels identify that SPACE/Early Help panels are operational and integrated into normal ways of working. A regional SPACE Wellbeing Steering Group has been established to enable governance and improvement. Outside the pending formal evaluation, some institutional factors in relation to roll-out have been identified (Personal communication 2022).

*Factors of relevance to innovation roll out across institutions in Wales:*

i. **Awareness of effort needed and time constraints for staff** in being involved (including concerns around demand, long waiting lists, insufficient contact time).
ii. **Constraints associated with short term funding** affecting planning and recruitment/retention of staff.
iii. **Need for** multi-agency involvement, liaison and communication.
iv. iv. Need for **changes to support implementation**; For this particular intervention the different referral forms in each borough and lack of clarity around services represented on each panel were identified as barriers.

#### Innovations in Wales with no local evaluation as yet

##### 5: Family Voice (Family Group Conferencing)

Family Group Conferencing is a conferencing model for families on the brink of court proceedings, aiming to place children within their family network. In Wales, this is called ‘Family Voice’ and is currently being evaluated in Wales by CASCADE and due for completion in October 2025.

Family Group Conferencing was one of the many innovations evaluated within England as part of the Children’s social care innovation programme (Sebba et al. 2017a) and overall guidance for institutional innovation, from those innovations is summarised in Section 2.2 The evaluation of Family Group Conferencing in Darlington, England (where it is called ‘Family Valued’) (Collyer et al. 2021, included in Sebba et al. 2017a) has recommendations for introducing the intervention into a new LA including:

Training and support to achieve and maintain whole system change, key local roles to support and champion roll out and ongoing local leadership, clear communication and support for how new innovation will be integrated alongside other practice models, sustainable funding, longer term monitoring and evaluation (Collyer et al. 2021).

*Factors of relevance to innovation roll out across institutions in Wales:*

None identified for Wales. Welsh LAs might consider how far findings relating to adoption/scale-up of Family Group Conferencing in England may be applicable to the context of the Welsh implementation, or whether a local evaluation may be appropriate and feasible.

##### 6: The Mockingbird Family Model (MFM)

The Mockingbird Family Model (MFM) is a foster care delivery model that creates an extended family network to support, develop and retain quality foster families so they can meet the challenging and complex needs of children and young people in foster care. The MFM approach is to create clusters of 6-10 homes (satellites) to form a constellation which reflects the extended family structure and is supported by hub carers. As of 2021, there are 85 constellations in the UK including one in Flintshire in North Wales (2021 programme update). Flintshire is expanding the programme and will be evaluating its implementation.^4^

The MFM was one of the many innovations evaluated within England (McDermid et al. 2016, Ott et al. 2020) as part of the Children’s Social Care Innovation programme (Sebba et al. 2017a, FitzSimons et al. 2020).

Two evaluations, relating to Mockingbird roll-out in England (McDermid et al. 2016, Ott et al. 2020; included in Sebba et al. 2017a and FitzSimons et al. 2020) have findings relevant to the support of organisational innovation including:

Clear operating guidelines, clarity around information sharing (including safe-guarding), laying the groundwork and assuring buy-in from leadership, staff retention and funding sustainability (McDermid et al. 2016, Ott et al. 2020).

*Factors of relevance to innovation roll out across institutions in Wales:*

None identified for Wales. Welsh LAs might consider how far findings relating to adoption/scale-up of Mockingbird in England may be applicable to the context of the Welsh implementation in Flintshire and elsewhere, or whether a local evaluation may be appropriate and feasible.

##### 7: Signs of Safety (SoS)

Signs of Safety is a strengths-based, safety-organised approach to collaborative child protection that emphasises the central role of the relationship between the social worker and the parents. SoS has been evaluated in a systematic review (Sheehan et al. 2018) with an emphasis on studies from the UK and other relevant countries; 38 publications in all and 13 from the UK. One of the UK studies was based in Wales (City and County of Swansea, 2014) but the Swansea report did not explore institutional factors influencing innovation. Whilst the review (Sheehan et al. 2018) did not find firm evidence of efficacy in terms of reducing the need for children to be in care, authors found that SoS can lead to positive engagement across families and external organisations, and a number of institutional factors relating to innovation roll out were identified. These included:

Compatible data recording systems, used within organisations, creating a safe environment for honest and open feedback (an organisational learning culture), active leadership support and allowing the time required for multi-level organisational change.

*Factors of relevance to innovation roll out across institutions in Wales:*

None identified for Wales. Welsh LAs might consider how far findings relating to the adoption/scale-up of SoS in the UK and similar countries may be applicable to the context of the Welsh implementation in Swansea and elsewhere, or whether a local evaluation may be appropriate and feasible.

### 2.3 Bottom line results

A number of factors have been identified that may affect the scale and spread of innovations in children’s social care and have been outlined above (Sections 2.1 & 2.2). The identified factors across the body of evidence are summarised below in Table 1 and are listed in more detail in Section 6.2.

**Table 1:**
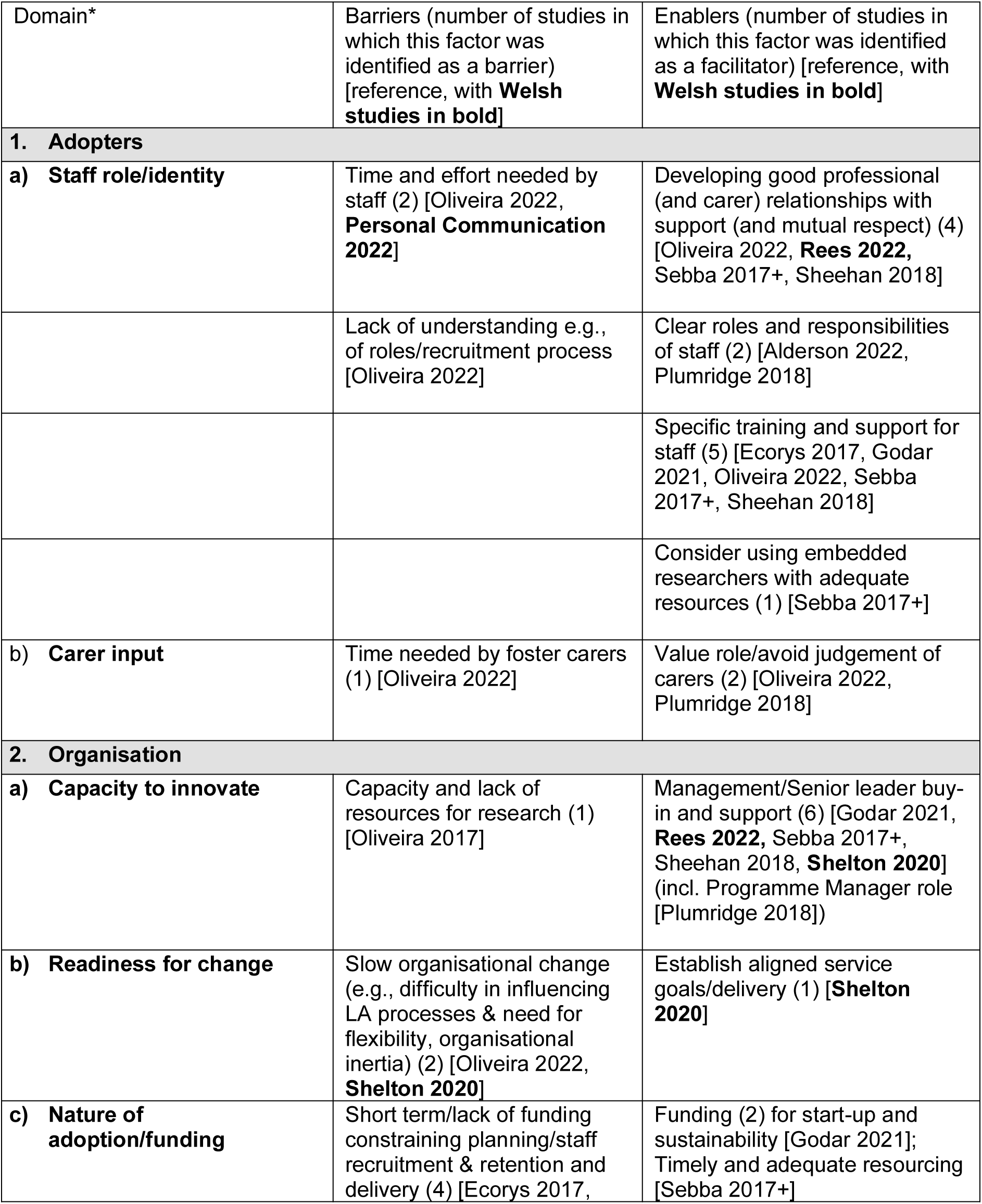
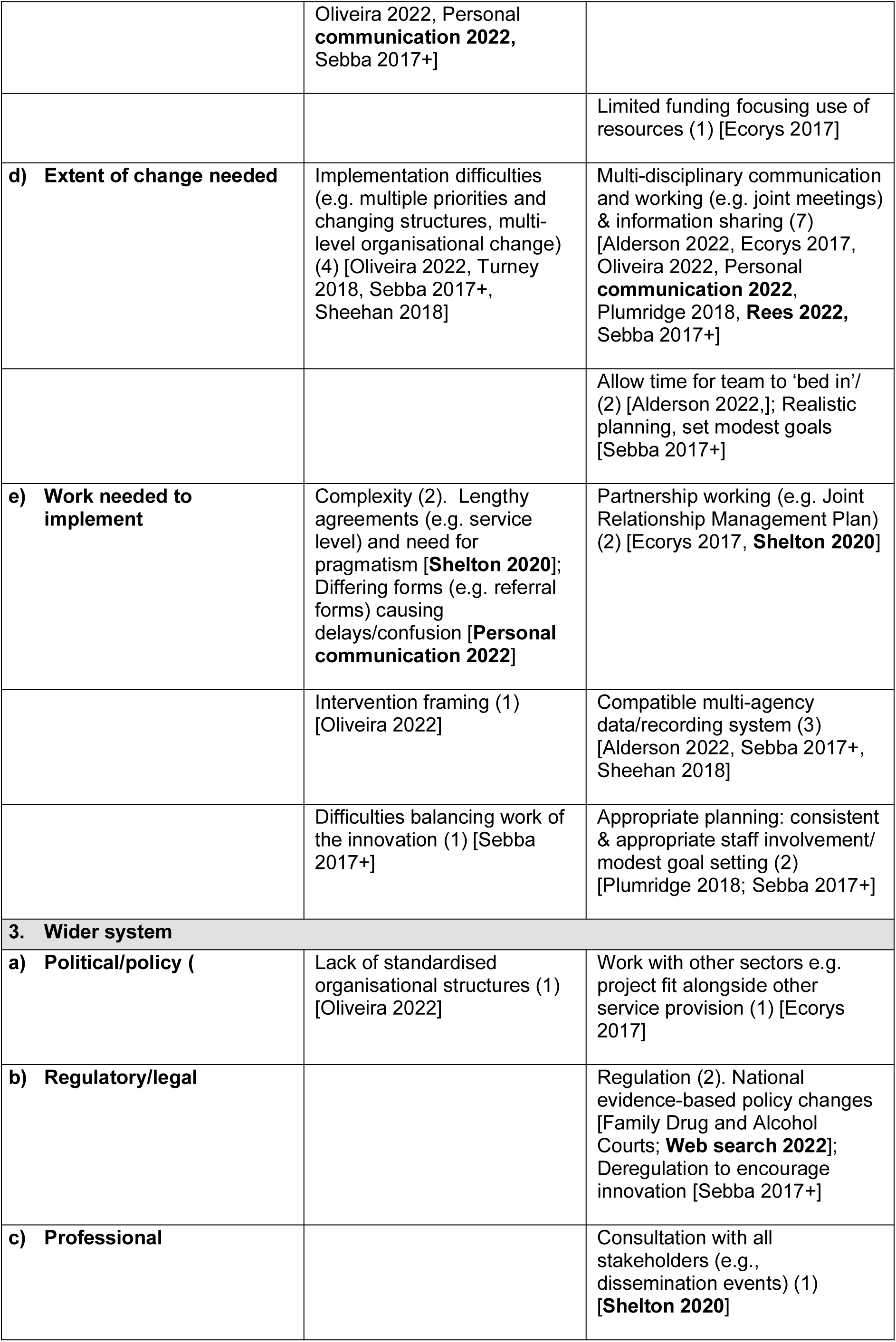
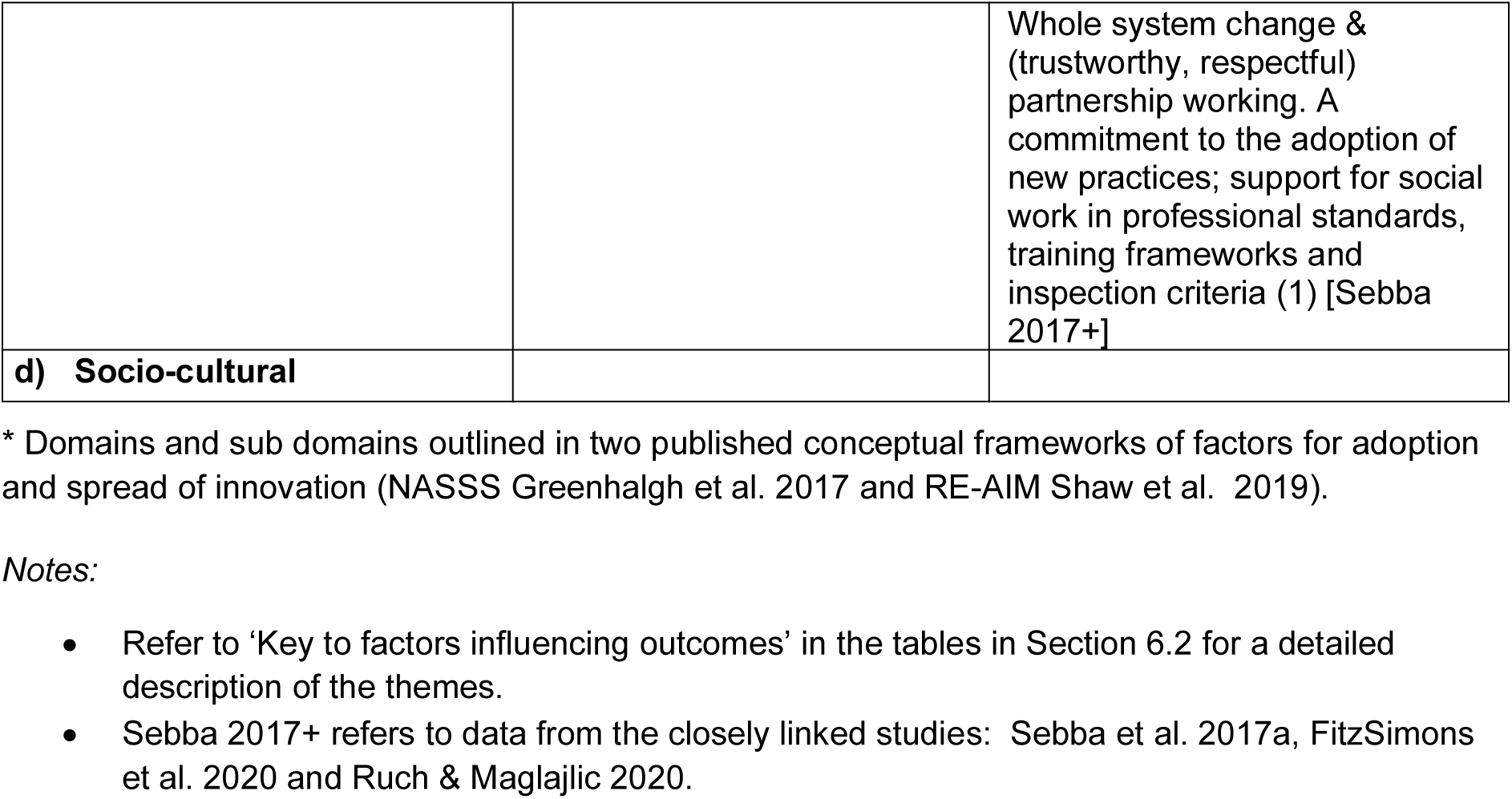
Organisational factors that may act as barriers or enablers to the scale and spread of innovations in children’s social care in the UK.

The study designs included non-systematic reviews and service evaluations for which there is no published critical appraisal form. Formal critical appraisal was carried out in five out of the ten included studies. All of the studies had some quality limitations.

## 3. DISCUSSION

### 3.1 Summary of the findings

There are some organisational facilitators relating to the scale and spread of innovations in children’s social care for which a clear consensus seems to be emerging across this diverse body of literature:

Specific training and support for professional staff
Support and mutual respect within inter-professional and professional-carer relationships
Senior management/leadership buy-in and support
Multi-disciplinary communication and joint working
Develop compatible data systems to support joint working/collaboration

Barriers for which a clear consensus seems to be emerging for the literature were:

Short term or lack of funding (the need for funding was presented as an enabler in some studies)
Implementation difficulties (e.g., organisational inertia, multiple priorities and changing structures)

The number of studies that report each theme are presented in Table 1 (Section 2.3) but should not be equated directly with a hierarchical order of importance. There are other factors that should also influence how much confidence a reader might have in each finding (e.g., the design and quality of the contributing studies and the relevance of the populations and settings to ones’ own context).

From personal contacts and web searching carried out for this review, it is clear that there is a huge variation and complexity of organisational structures across Wales that may, of themselves, act as barriers to innovation across authorities.

### 3.2 Strengths and limitations of the available evidence

This type of review doesn’t lend itself to the rapid approach required by the time frame.

Included studies were a mixture of those designed to look at innovation scale/spread and those looking at a specific intervention but with innovation relevant findings. For the latter it was sometimes difficult to tease out the innovation-specific recommendations from the intervention-specific ones
All included studies had quality limitations.

### 3.3 Implications for policy and practice

This rapid review highlights the complexity of the social care models but provides some clear pointers for policy and practice. The findings are stated above (Section 3.1) and all have direct implications for policy and practice.

*Comparison with research in other sectors:*

The findings from this review align well with those from the adult social care reviews recommended by stakeholders; the Supporting Adult Social Care Innovation (SASCI) report (Zigante et al. 2022) and the Social Care Institute for Excellence (SCIE) report (Callanan & Mitchell 2020).

The findings from this review align to some extent with all five of the themes noted by Zigante et al. (2022) and three noted by Callanan & Mitchell (2020). This review identified:

**Senior management buy-in and support.** Zigante et al. (2022) highlighted leadership and Callanan & Mitchell (2020) support from leadership and management.
**Short and longer term funding.** Zigante et al. (2022) highlighted resources (financial and human) and Callanan & Mitchell (2020) the need for funding.
**Multi-disciplinary communication and joint working**. Zigante et al. (2022) highlighted collaboration and Callanan & Mitchell (2020) networks for spreading innovations and sharing knowledge.
Good professional (and professional-carer) relationships with support and mutual respect. Zigante et al. (2022) highlighted culture.
**Specific training and support for professional staff**. Zigante et al. (2022) highlighted knowledge and evidence.

Other findings from Callanan & Mitchell 2020 (for factors external to the innovation) included acknowledging the inherently difficult environment, need to allow time for scaling and embedding, and the use of policy and financial levers to encourage adoption. These issues were also touched on within this review.

Where innovations have not yet been formally evaluated within Wales it may be worth considering whether the context in Wales is sufficiently different to require formal evaluation in relation to guidance for institutions in scaling innovation in children’s social care.

### 3.4 Strengths and limitations of this Rapid Review

This review was conducted rapidly to inform policy and decision makers, and therefore methods were adjusted as an understanding of the evidence base developed. A comprehensive search strategy was designed to identify relevant evidence in the bibliographic databases. In addition to the databases, we searched grey literature and screened publications highlighted by the stakeholders, as being potentially relevant. Within this review we adopted an inclusive approach and therefore also included examples relevant to policy of best practice interventions implemented in Wales. This review has a strong reliance upon grey literature and overview reports consisting of evaluations undertaken across UK. The combination of implementation examples, overviews and implementation or process evaluations can contribute new understandings and identify factors that support or inhibit the scale and spread of innovations in children’s social care.

The time frame of the review precluded a methodologically robust thematic analysis. The authors of this review attempted to match findings to the most appropriate theme(s) but some of these could fit into two or more themes and there is potential for overlap. The approach was to focus on primary studies and articles relating to implementation or process evaluations combined with examples of interventions implemented in Wales. This review benefitted from two authors (rather than a single author) matching factors identified from the studies to the Framework in 2. A formal thematic analysis (using specialist software e.g. NVivo) would have been ideal had time permitted.

In conducting this review rapidly, it should be noted that data extraction and critical appraisal of each study were undertaken by different reviewers although they were independently checked for accuracy and consistency. Also, studies were included regardless of their quality.

If time had allowed, the ideal would have been a global view (relevant settings) of innovation in children’s social care leading to an agreed conceptual framework before drilling down to UK and Wales.

The framework of relevant factors was adapted from two published frameworks of factors for organisations adopting/scaling up innovation (NASSS and RE-AIM) and then adapted for the factors identified within this review, rather than being developed specifically from the global overview approach outlined above.

## Data Availability

All data produced in the present study are available upon reasonable request to the authors

## 5. RAPID REVIEW METHODS

### 5.1 Eligibility criteria

**Table 2:** Eligibility Criteria.

5.1 Eligibility criteria.
| Table 2: Eligibility Criteria |  |  |
| --- | --- | --- |
|  | Inclusion criteria | Exclusion criteria |
| **Population** | Children's social care organisations (including statutory organisations and third sector) | Healthcare |
| **Intervention / exposure** | Innovation - For the purpose of this review, innovation will be defined as: *"Innovation is the implementation in practice of an idea, practice or invention within an organisation or system that is novel to the organisation or system. The introduction of the new idea, practice or invention produces a process of change with uncertain outcomes that is disruptive for the individuals, organisation, or system."* SASCI (Supporting Adult Social Care Innovation) Project, led by the LSE. |  |
| **Comparison** | Current practice, another comparable intervention, or no comparator |  |
| **Outcome measures** | Primary outcomes:<br>All organisational factors (barriers and enablers) that affect the implementation and scale up of an innovation.<br>Secondary outcomes:<br>Any quantitative measures of the adoption (in one setting), spread (replication in another setting) and scale-up (building infrastructure to support full scale implementation) of innovation. | Articles describing the sharing of information on an innovation but with no data on institutional factors relating to adoption or scale up. |
| **Setting** | Children's social care <18 years old | Adult social care >18 years old. |
| **Context** | Responses to social need in children and families, including family support services, children in need services, child protection, looked after children and adoption. This also includes advocacy services and care and support for daily living provided to disabled children and their families. |  |
| **Study design** | Any UK based primary-research study design (qualitative, process evaluation etc.) including individual | Editorials, conference abstracts, opinion pieces. |
|  | case studies and secondary-research with specific relevance to the UK. |  |
| **Countries** | United Kingdom | Rest of the world |
| **Language of publication** | English |  |
| **Publication date** | 2014 - 2022 | Pre-2014 |
| **Publication type** | Published, Unpublished and preprint |  |
| **Other factors**<br>*Any other key points to note* |  |  |

NOTE: The review adopts a broad definition of innovation:
“Innovation is the implementation in practice of an idea, practice or invention within an organisation or system that is novel to the organisation or system. The introduction of the new idea, practice or invention produces a process of change with uncertain outcomes that is disruptive for the individuals, organisation or system.”

### 5.2 Literature search

Prior to planning this review, a preliminary review of existing reviews was conducted. The findings were presented to the stakeholders and used to refine the scope of the present rapid review of primary studies, and to inform the methods. For details of all the resources searched, please refer to Appendix 1.

A comprehensive search was designed in Social Policy and Practice (see Appendix 2) to identify relevant primary studies and was then translated to the databases listed in Appendix 1. It uses a combination of text words, social science thesaurus terms and medical subject headings. Known literature provided by stakeholders was also checked for eligibility and included or used as a source of specific relevant evidence.

The grey literature search consisted of reports identified by the review team or provided by Stakeholders. Additionally, a list of grey literature websites was provided by the stakeholders which were searched, along with websites identified by the review team. For searching grey literature resources, a broad search using word variations of the terms: innovation and implement* and scale and “children’s social care” were conducted.

Searching was completed on 18 November 2022.

### 5.3 Reference management

Database searches were imported into Endnote 20 and deduplicated by a single reviewer. Grey literature search results were added to an Excel spreadsheet and cross-checked against the Endnote library.

### 5.4 Study selection process

Two independent review authors carried out initial screening and exclusion for the identified titles and abstracts. The full-text study selection was conducted by individual reviewers. Eligibility criteria was used to assess the titles and abstracts and then full text of all sources identified by the search. Grey literature reports were also assessed for eligibility by individual reviewers. Where one reviewer was uncertain as to inclusion it was checked by a second reviewer. The reference lists of any identified systematic reviews were also scanned for any additional relevant primary research.

### 5.5 Data extraction

The NASSS (non-adoption, abandonment, scale-up, spread, sustainability) framework (Greenhalgh et al. 2017) identifies seven domains which relate to the innovation and its adoption or spread. We adapted the NASSS framework (Figure 1) and the five factors of RE-AIM (reach, effectiveness, adoption, implementation, maintenance) framework (https://re-aim.org/; Shaw et al. 2019) to develop a framework (see *Key to factors influencing outcomes* Tables 3-5) to capture the appropriate data to address the review question.

**Figure 1:**
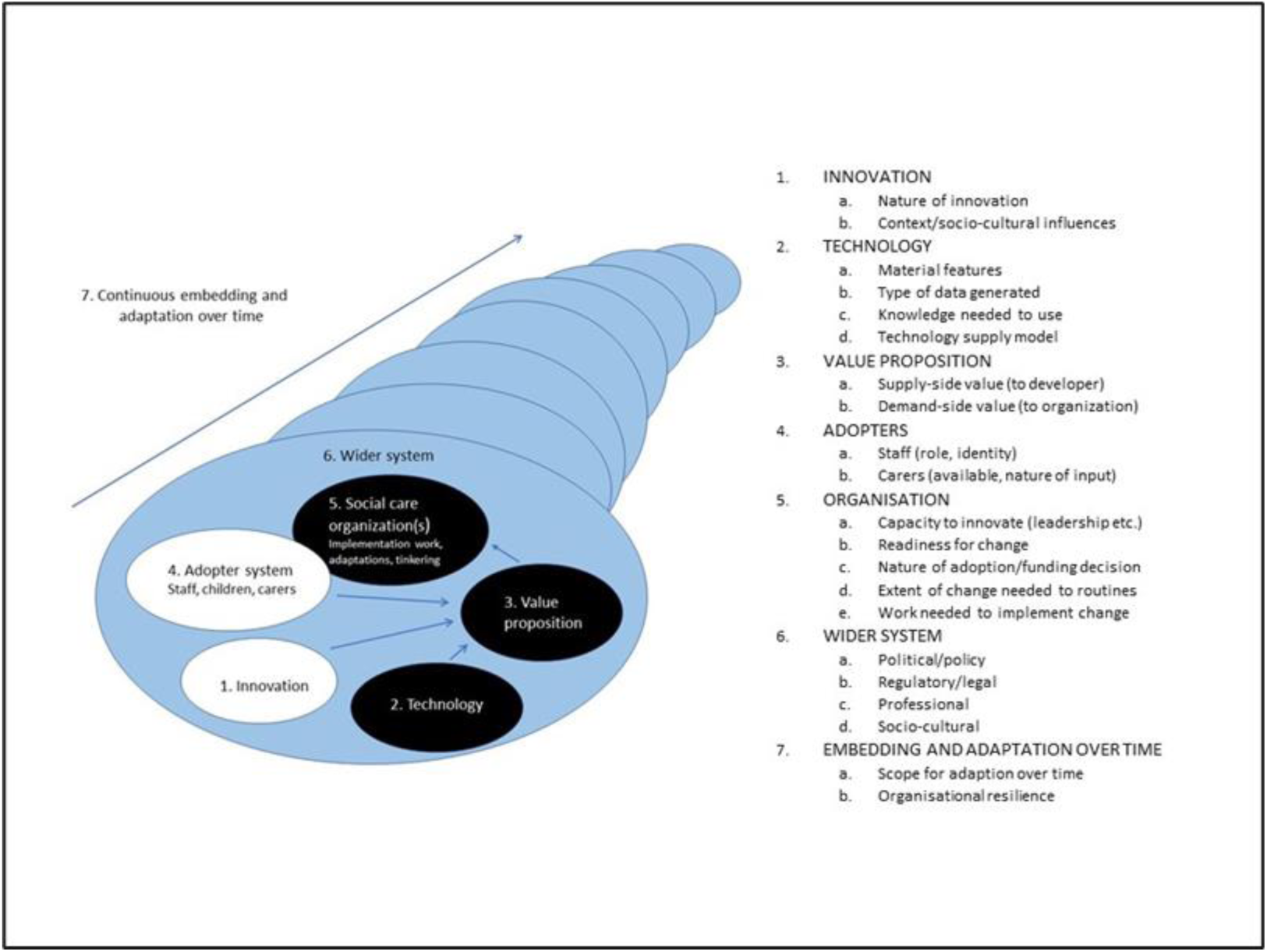
Framework of factors that may support or inhibit the scale and spread of innovations in children’s social care (adapted from Greenhalgh et al. 2017)

Data on factors potentially influencing scale and spread of innovation were extracted from studies and reports into the data extraction forms which also captured additional key information such as participants, outcomes investigated, evidence type, data collection methods data extraction was completed by individual reviewers and checked by a second reviewer (Section 6.2).

### 5.6 Quality appraisal

The methodological quality of included studies was assessed using the following critical appraisal tools:

AMSTAR 2: Critical appraisal tool for systematic reviews that include randomised or non-randomised studies of healthcare interventions, or both.
CASP Qualitative Studies Checklist

Papers relating to implementation or process evaluations were included regardless of their quality but have been commented on within the narrative summaries. A pragmatic system has been devised and reported to assess each study as high, medium or low quality for the purposes of this rapid review.

Quality assessments were completed by a single reviewer and any uncertainties were checked by a second reviewer. Notes on study quality are recorded in the summary tables (Tables 3-5, Section 6.2) and all quality assessment data are available from the authors.

### 5.7 Synthesis

The findings of this review are presented narratively. Data from the included studies are summarised and presented in tables. For this review we have adopted an inclusive approach, using a wide range of evidence relevant to policy to draw new understandings and identify factors that support or inhibit the scale and spread of innovations in children’s social care.

## 6. EVIDENCE

### 6.1 Study selection flow chart

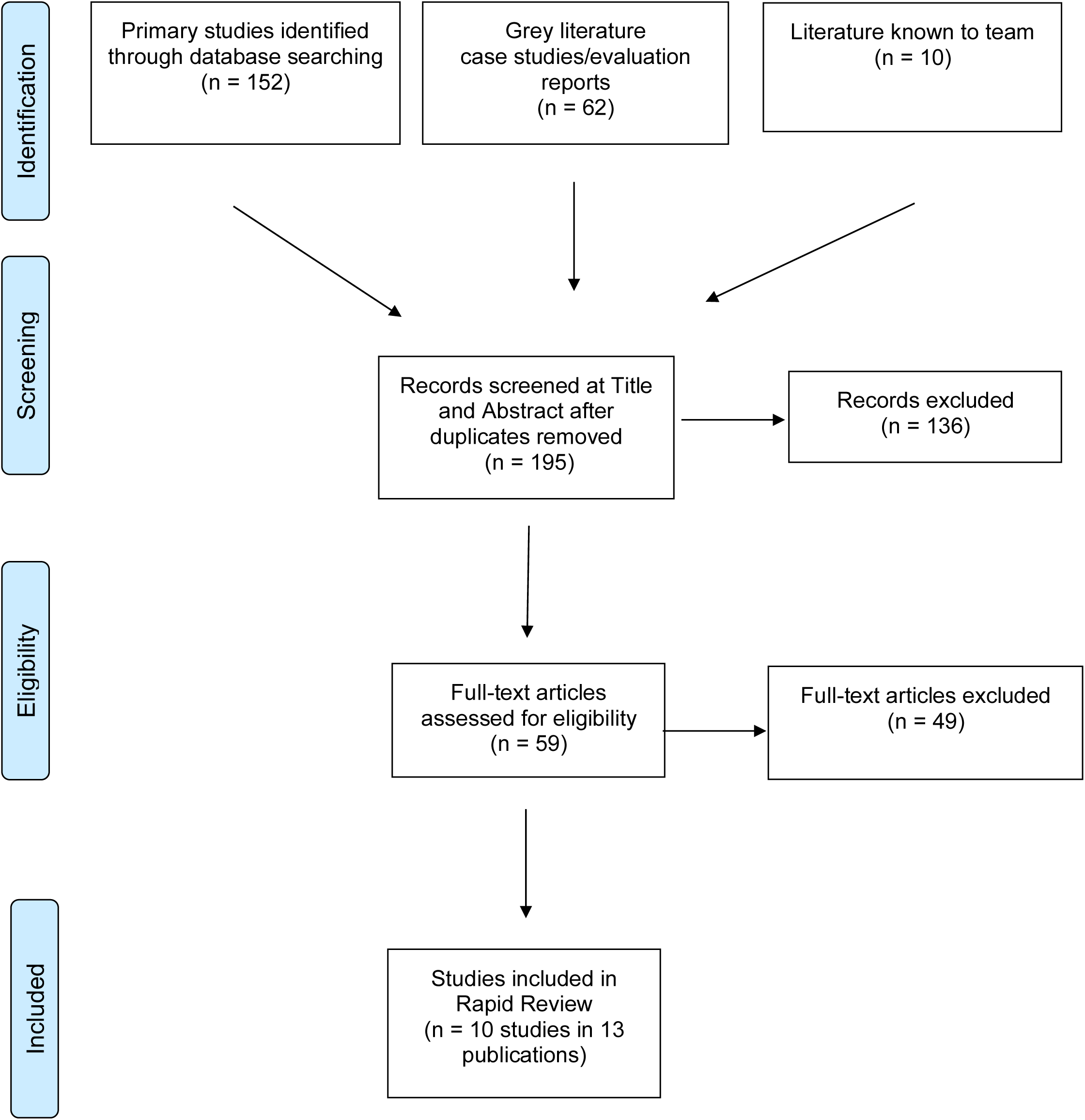

### 6.2 Data extraction tables

**Table 3:**
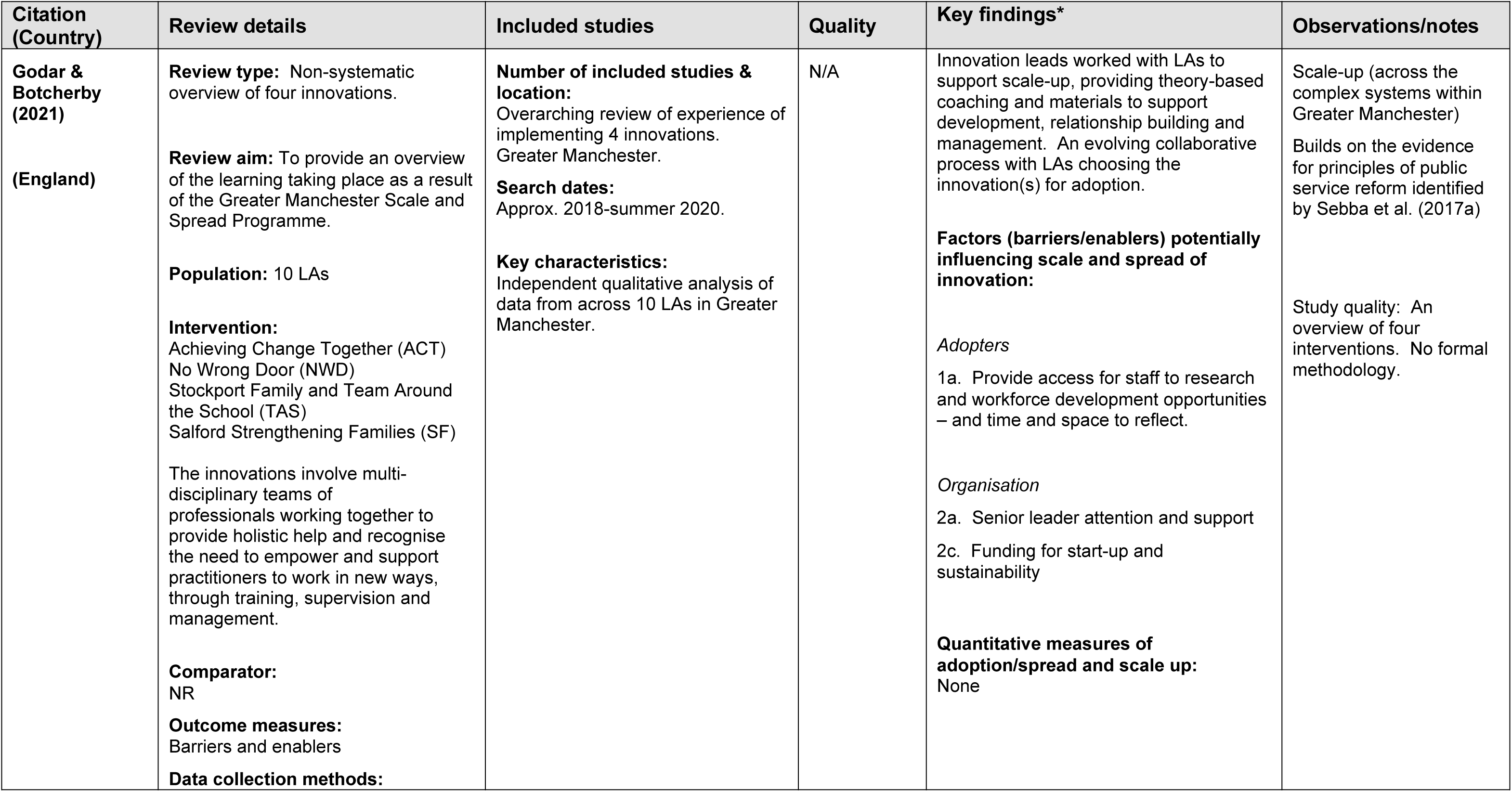
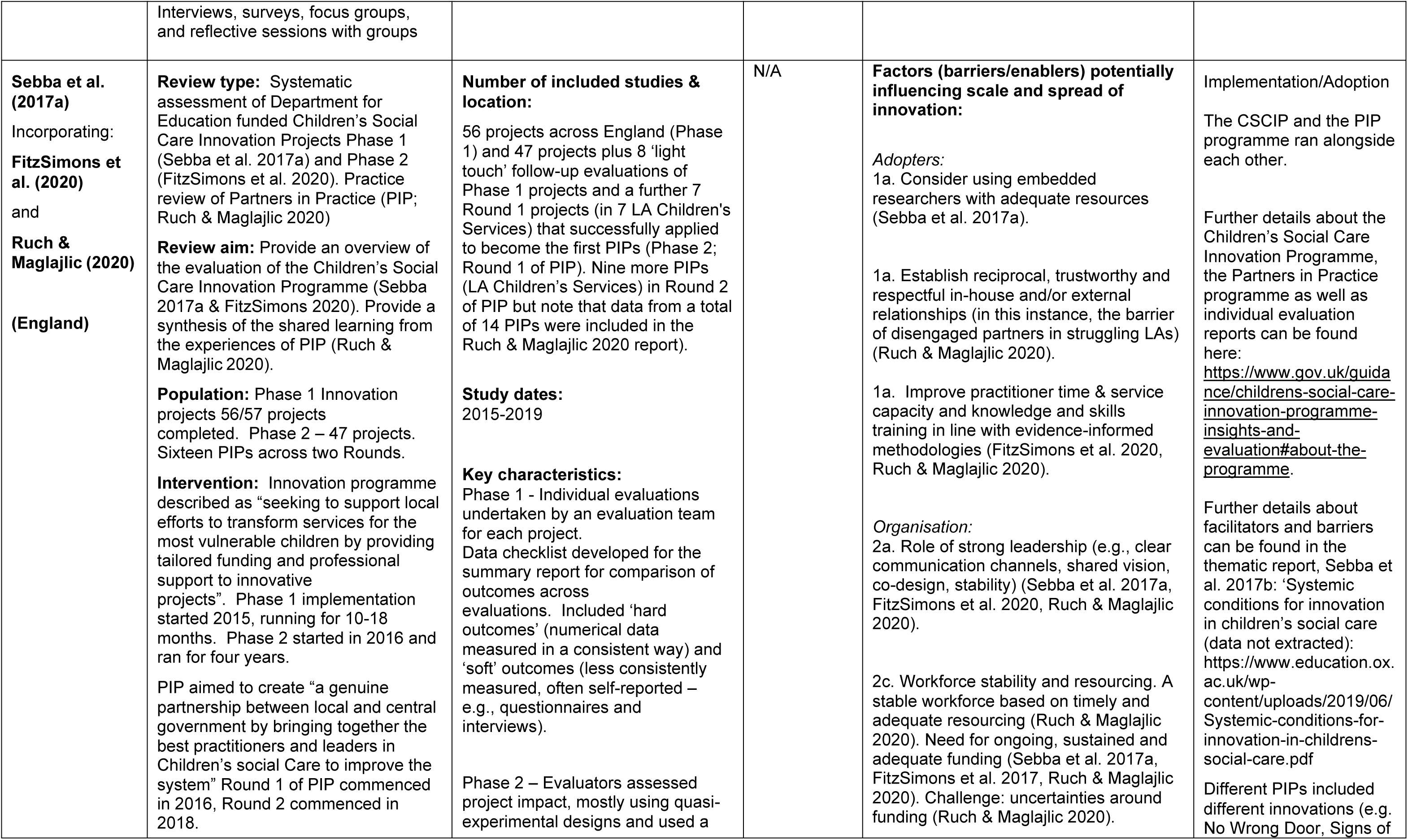
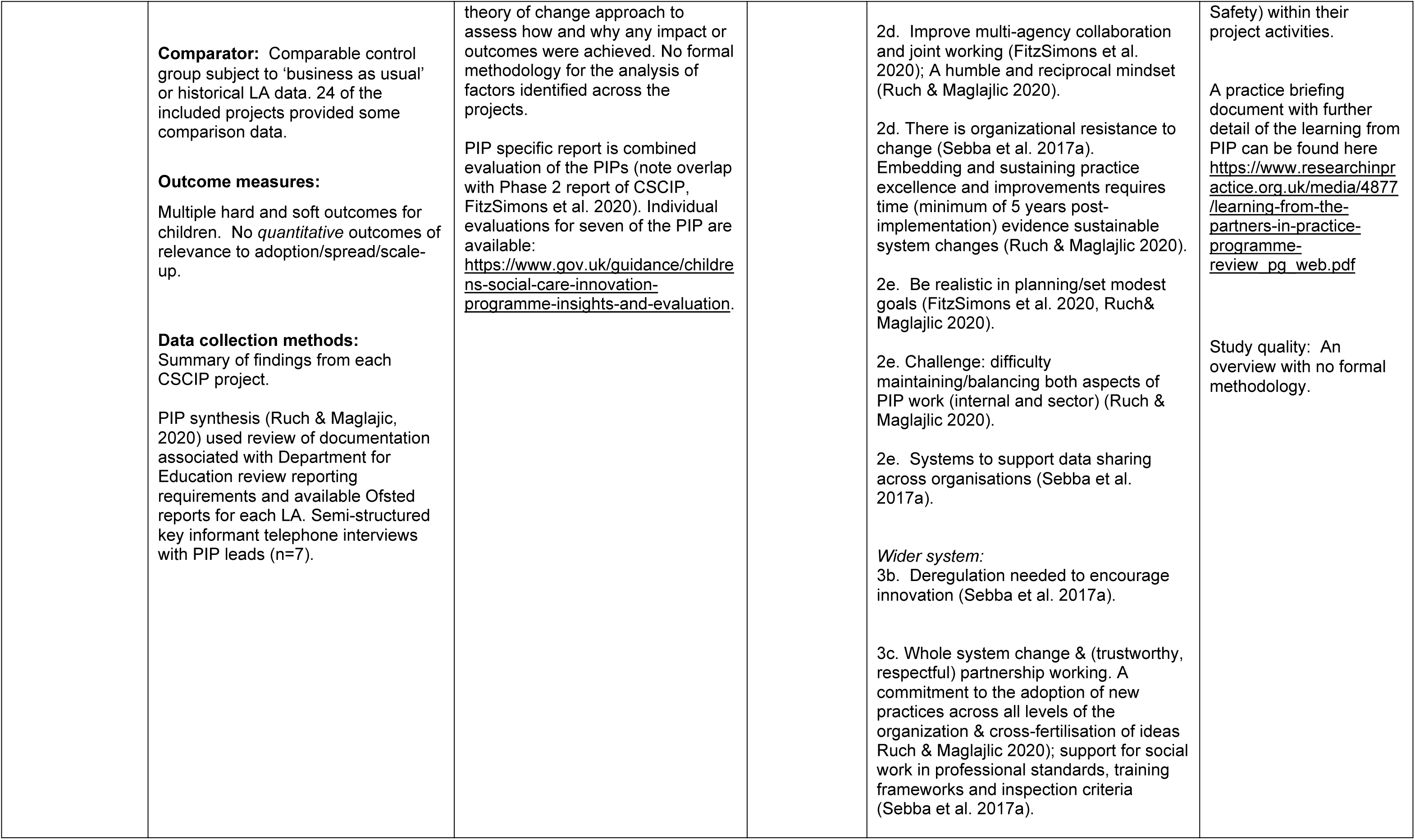
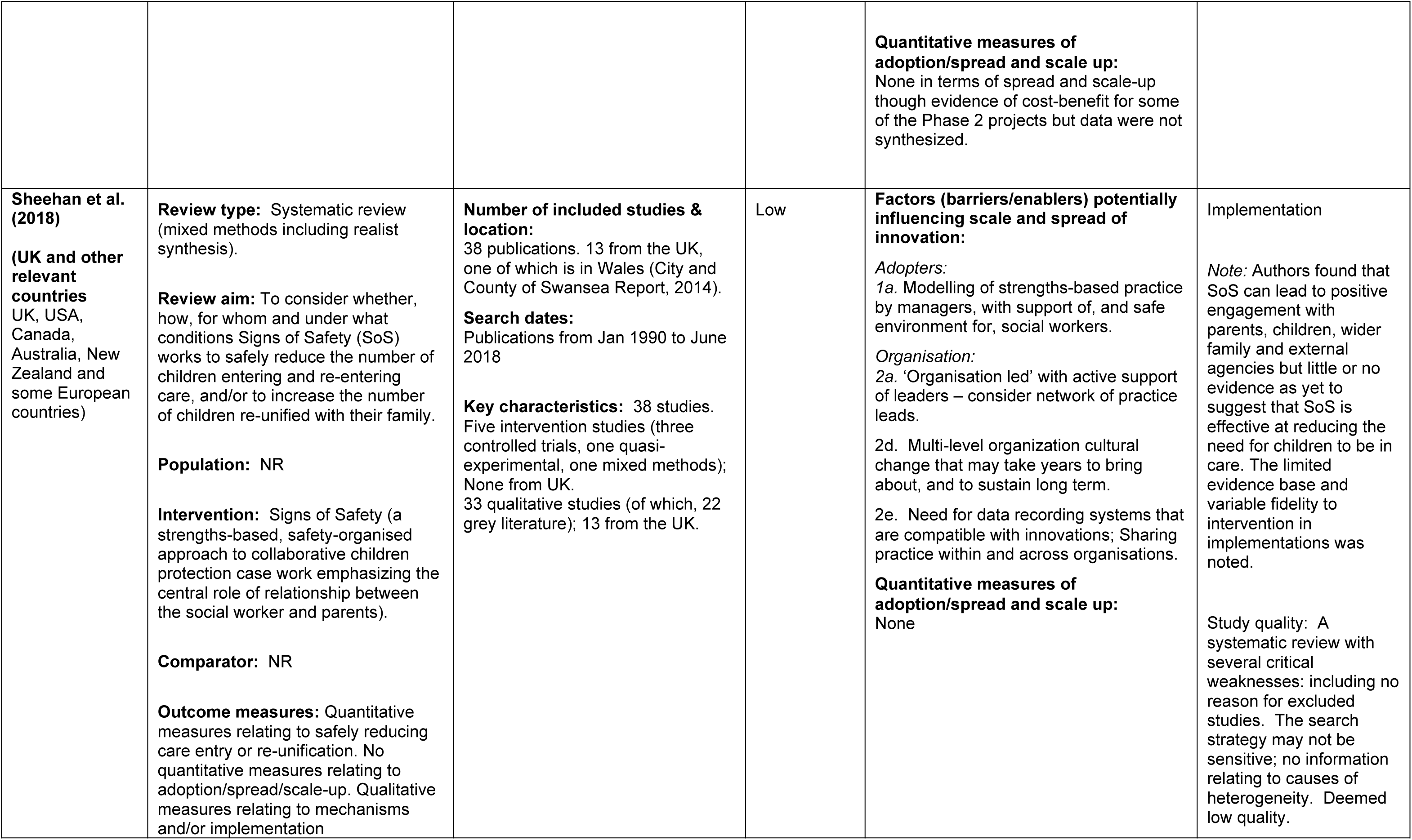
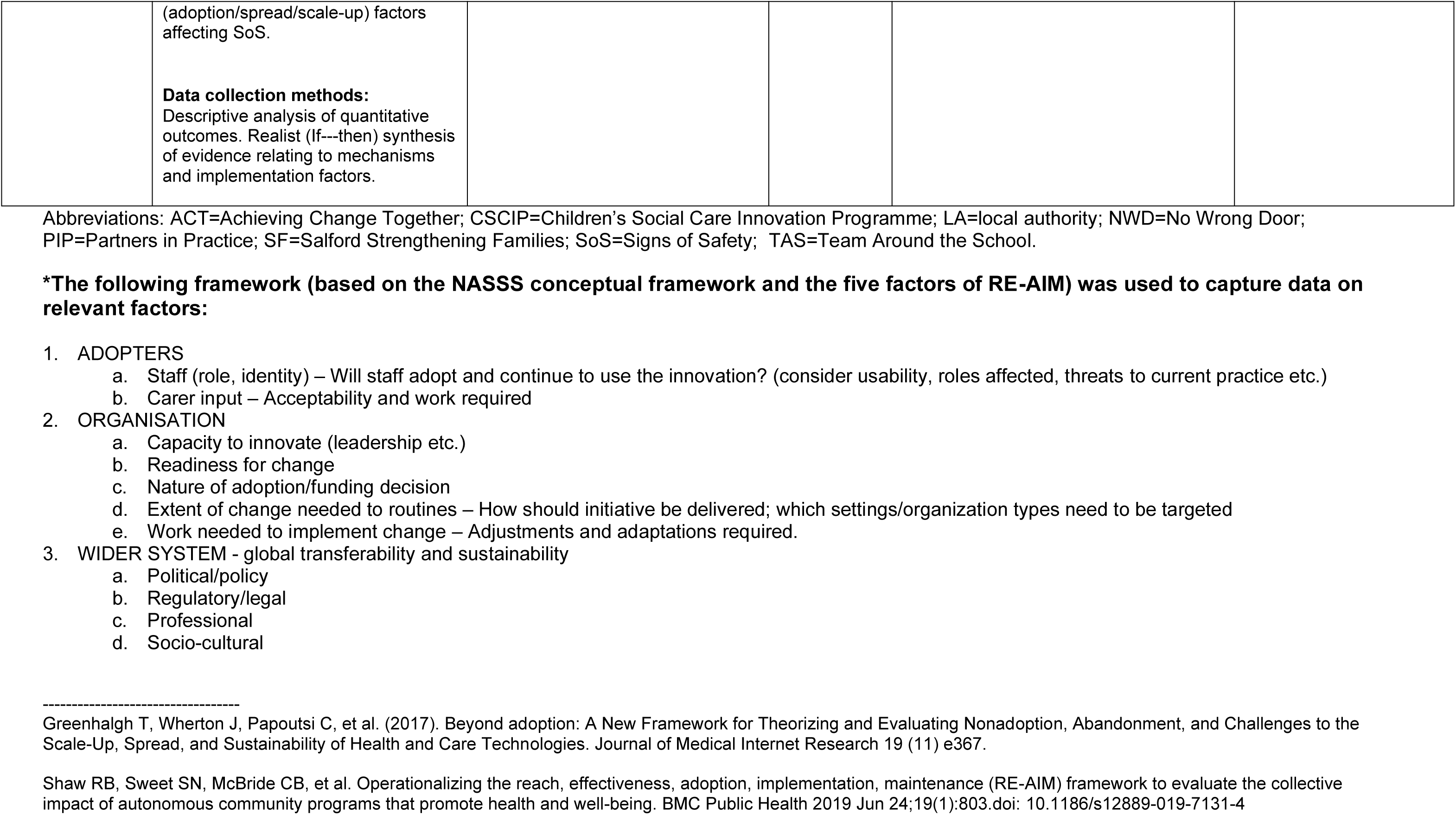
Summary of secondary research with specific relevance to the UK.

**Table 4:** Summary of primary research in the UK (excluding Wales)

| Citation (Country) | Study Details | Participants & setting | Quality | Key findings* | Observations/notes |
| --- | --- | --- | --- | --- | --- |
| Alderson et al. (2022)<br>(England) | **Study Design:** Qualitative<br>**Study aim:** examine the implementation, service delivery and perceived impact for family members of the innovation.<br>**Project name:** Innovation Pilot Project, IPP<br>**Project aim:** To reduce fragmentation between services and help identify children affected by parental alcohol misuse.<br>**Type of innovation:** Communication and collaboration across child welfare systems<br>**Data collection methods:** Qualitative including interviews with 27 family members and interviews/focus groups with 14 staff | **Geographic location:** Newcastle, England<br>**Lead authority/organisation:** Newcastle City Council<br>**Setting:** Multi-agency collaboration<br>**Dates of data collection:** January 2020 – October 2021 | Medium | **Factors (barriers/enablers) potentially influencing scale and-spread of innovation:**<br>*Adopters:*<br>1a. Clear roles and responsibilities of staff including appointment of project coordinator.<br>*Organisation:*<br>2d. Allow time for team to 'bed in' (joint meetings).<br>2e. Establish multi-agency recording system.<br>**Quantitative measures of adoption/spread and scale up:**<br>None | Implementation<br>Study quality: No information on recruitment; relationship between researcher and participants not adequately considered.<br>Authors concluded keeping with the previous literature was a challenge. Due to COVID-19 restrictions service delivery was affected in terms of how interventions were delivered and access to interviewees as part of the study.<br>The authors reported that in the beginning, supervision was conducted by their original employing organisations. That was regarded to be problematic due to the project workers day-to-day work being overseen by a manager within the IPP team.<br>Deemed to be medium quality |
| Ecorys UK (2017)<br>(Scotland) | **Study Design:** Service Evaluation<br>**Study aim:** To evaluate the Dundee Early Intervention Team's Improving Futures project. (The report provides a profile of families support, main lessons learned, main | **Geographic location:** Dundee<br>**Lead authority/organisation:** Aberlour Child Care Trust<br>**Setting:** Voluntary and Community Sector | N/A | **Factors (barriers/enablers) potentially influencing scale and-spread of innovation:**<br>*Adopters:*<br>1a. Intervention specific training for staff important – In this instance social pedagogy training to support relationship building with families). | Implementation. Part of the Improving Futures project (26 pilot projects across the UK to test different Voluntary and Community Sector led approaches).<br>Barriers and enablers extracted from within the chapters on lessons learned from project delivery and on |
|  | achievements, overall conclusions and recommendations).<br>**Project name:** Dundee Early Intervention Team's Improving Futures project.<br>**Project aim:** to establish an early intervention and preventative support service for families in Dundee who did not meet the threshold for statutory intervention. To work with families to find and make effective and sustainable solutions to social, health and relationship or parenting difficulties, both before and at the point of crisis.<br>**Type of innovation:** Early intervention and preventative service for children and families to strengthen and complement Dundee's Integrated Children's Services offer. Delivered by a partnership of organisations: Aberlour Child Care Trust, Barnardo's Scotland, CHILDREN 1<sup>st</sup> and Action for Children Scotland. Key aspects included:<br>1) Support for those who do not attend health appointments.<br>2) Support for families who don't attend any support groups (limiting social interaction & social development). | **Dates of data collection:** Funding for project awarded Jan 2012. Qualitative case study visit in November 2014. |  | **Organisation:**<br>2c. Short-term funding created challenges. In particular to divert resources to focus on sustainability of the service and effects on staff changes. However, diverting resources also led to changes in the referral and allocation process which led to a more effective service with families most in need being supported.<br>2e. Partnership working. Operational group involving staff from each partner organisation shared learning and information.<br>**Wider system:**<br>3a/c. Working with other services including Social Work, Education and Health to ensure project fit alongside other service provision.<br>**Quantitative measures of adoption/spread and scale up:**<br>None | main achievements and sustainability of the project.<br>Study quality: A service evaluation with no formal methodology |
|  | 3) Lower threshold concerns e.g., healthy food choices.<br>4) Provision of practical support e.g., managing the after nursery/school period).<br>**Data collection methods:**<br>1) desk review of documentation; 2) project monitoring data inputted by project staff (154 families at project entry point); 3) qualitative case study visit: interviews with staff, stakeholders and families; 4) in-depth interview with project co-ordinator. |  |  |  |  |
| **Oliveira et al. (2022)**<br>**(England)** | **Study Design:**<br>Qualitative study<br>**Study aim:** To understand the impact, acceptability and feasibility of implementing a trial of the Video-feedback Intervention to promote Positive Parenting and Sensitive Discipline, Foster Care (VIPP-FC) in a CAMHS and LA context. To understand the facilitators of and barriers to the implementation of the VIPP-FC.<br>**Project name:**<br>VIPP-FC (UK): Video-feedback Intervention to | **Geographic location:**<br>Sites included urban (Greater London, Peterborough) and rural/semirural (Yorkshire/Hertfordshire) areas.<br>**Lead authority/organisation:**<br>Outpatient NHS mental health services across eight trusts and nine partner social services departments.<br>**Setting:**<br>Outpatient child and adolescent mental health services (CAMHS) and partner social services departments. Sites included urban and rural/semirural areas. | Medium | **Factors (barriers/enablers) potentially influencing scale and-spread of innovation:**<br>*Adopters:*<br>1a. the study was not a mandatory activity and involved extra work for some staff, so could be easily placed on the 'backburner'.<br>1a. Giving time and resources to building relationships with all the key players.<br>1a. Lack of understanding of the recruitment process (including roles), set-up and delivery as well as study aims, approach and benefits.<br>1a. Staff learning from intervention training brought new perspectives to their work. Opportunity for staff to be offered continued professional development.<br>1b. It was noted that foster carers can sometimes feel 'judged' by professionals, so they emphasised the | This is an adaptation of an intervention from the Netherlands (adoption) which is then feasibility and piloted tested (so assuming "implementation" at that point).<br>Details extracted here are from Chapter 4, a qualitative study which is part of a wider mixed methods study adapting the intervention and conducting a feasibility study and pilot RCT to inform a full-scale RCT.<br>Study quality: A reasonably well conducted study deemed medium quality. Appropriate sample but unclear whether *all* relevant stakeholders were invited to participate. Lack of consideration of |
|  | promote Positive Parenting and Sensitive Discipline, Foster Care<br>**Project aim:** To promote sensitive parenting and secure attachments in young children in foster care with reactive attachment disorder symptoms. It aims to: 1) improve sensitive responding of foster carer, 2) improve consistent responding of foster carers to challenging child behaviour, 3) improve foster carer-child relationships and 4) improve reactive attachment symptoms and the child's emotional and behavioural outcomes.<br>**Type of innovation:** The Video-feedback Intervention. Practitioner films the child and carer interacting and provides feedback in the following session.<br>**Data collection methods:** Semi-structured interview schedule which lasted 50–70 minutes and were audio-recorded. Two of the interviews were carried out by telephone because of a lack of availability for face-to-face meetings. | **Dates of data collection:** Qualitative study January and March 2020. |  | importance of being clear about the potential benefits to foster carers and specifically emphasising its strengths-based approach.<br>1b. Difficulties faced with the available time of foster carers.<br>*Organisation:*<br>2a. Capacity and lack of resources for research, primarily in the LAs.<br>2b. The challenges of change and the organisational inertia that one often faces when introducing something new within LAs.<br>2d. Difficulties implementing a study within the professional network such as managing multiple priorities and the often-changing structures within LAs.<br>2d. Good face-to-face communication needed (e.g., network meetings and joint working).<br>2d. Sharing of information and learning between the research team and LAs, and across the entire network.<br>2e. Consider framing the intervention - in this case as a 'training opportunity' for foster carers rather than 'attachment based'.<br>*Wider system:*<br>3a. Lack of standardised organisational structures across different LAs a barrier to research.<br>**Quantitative measures of adoption/spread and scale up:**<br>None | relationship between researcher and participants. |
| Plumridge & Sebba (2018) | **Study Design:** Mixed methods service evaluation. | **Geographic location:** Birmingham | N/A | **Factors (barriers/enablers) potentially influencing scale and-spread of innovation:** | Implementation. |
| (England) | **Study aim:** to investigate whether, and how, the project supports the young person in moving from a residential home to a stable foster care placement successfully for a minimum of a year. It also considered what works well and less well in making, stabilising and sustaining this move and what outcomes are achieved for young people.<br>**Project name:** Step Down Programme.<br>**Project aim:** to bring young people out of residential homes into foster placements. A key outcome is placement stability after 52 weeks.<br>**Type of innovation:** underpinned by a Social Impact Bond contract, funded by a social investor (Bridges Ventures) who pays the additional service cost on top of what the Council can meet. They then receive payment back if the young person stays in a placement for 52 weeks. The programme involves a carefully planned matching process and high levels of support including planned respite care. | **Lead authority/organisation:** Birmingham City Council (the commissioners) and Core Assets (the delivery organisation)<br>**Setting:** Transition to foster care.<br>**Dates of data collection:** Placements made Nov 2014 to Apr 2018. |  | **Adopters:**<br>1a. Role of staff (e.g., mentor, therapist) in supporting carer/young person.<br>1b. Value the role of and support carers.<br>**Organisation:**<br>2a. Programme Manager role - increased involvement beneficial for consistency between placements and links between placements, social workers and managers.<br>2d. Proactive use of 'progress meetings' linked to positive outcomes.<br>2e. Involvement of a consistent LA social worker who knew the child and ideally their family was helpful.<br>**Quantitative measures of adoption/spread and scale up:**<br>None | **Study quality:** A service evaluation with no formal methodology |
|  | **Data collection methods:**<br>Quantitative data collected as part of service delivery (from 31 young people). Interviews (213, relating to 32 placements) with young people, their carers, children's and supervising social workers, and mentors. |  |  |  |  |
| Turney & Ruch (2018)<br>(England) | **Study Design:**<br>Qualitative<br>**Study aim:** To explore the contribution of an innovative approach to supervision to social work practitioners' assessment and decision-making practices<br>**Project name:**<br>The Cognitive and Affective Supervisory Approach (CASA)<br>**Project aim:** An innovative, practice-based method designed to try and enhance the quantity and quality of information available for decision-making by focusing on both the cognitive and affective dimensions of practitioners' experiences.<br>**Type of innovation:**<br>cognitive interviewing techniques originally designed to elicit best evidence from witnesses and victims of crime. | **Geographic location:**<br>England<br>**Lead authority/organisation:**<br>Social workers and their agencies, and the academic team<br>**Setting:**<br>Two LAs<br>**Dates of data collection:**<br>Not reported | Low | **Factors (barriers/enablers) potentially influencing scale and spread of innovation:**<br>*Organisation:*<br>2d. Introducing the innovation perceived as such a difficult task/process (in this example, the CASA, though benefits were noted for case planning)<br>**Quantitative measures of adoption/spread and scale up:**<br>None | This study is about adopting CASA individual commitment and a level of organisational re-thinking to promote a cultural shift in perception about the role and purpose of supervision.<br>A new approach to supervision. The project involved introducing a new framework for supervisory discussion into the day-to-day practice of social work supervisors.<br>Study quality: Poorly reported and lacked methodological detail; therefore, deemed to be low quality. |
|  | **Data collection methods:**<br>Interviews from 14 supervisors. Reflective discussion groups to explore their experiences. |  |  |  |  |

Abbreviations: CASA=Cognitive and Affective Supervisory Approach; CAMHS=Child and Adolescent Mental Health Services; LA=local authority N/A=Not applicable; NR=Not Reported; VIPP-FC=Video-feedback Intervention to promote Positive Parenting and Sensitive Discipline, Foster Care.
**\*The following framework (based on the NASSS conceptual framework and the five factors of RE-AIM) was used to capture data on relevant factors:**
1. ADOPTERS - a. Staff (role, identity) – Will staff adopt and continue to use the innovation? (Consider usability, roles affected, threats to current practice etc.) - b. Carer input – Acceptability and work required. 2. ORGANISATION - a. Capacity to innovate (leadership etc.) - b. Readiness for change - c. Nature of adoption/funding decision - d. Extent of change needed to routines – How should initiative be delivered; which settings/organization types need to be targeted - e. Work needed to implement change – Adjustments and adaptations required. 3. WIDER SYSTEM - global transferability and sustainability - a. Political/policy - b. Regulatory/legal - c. Professional - d. Socio-cultural

**Table 5:**
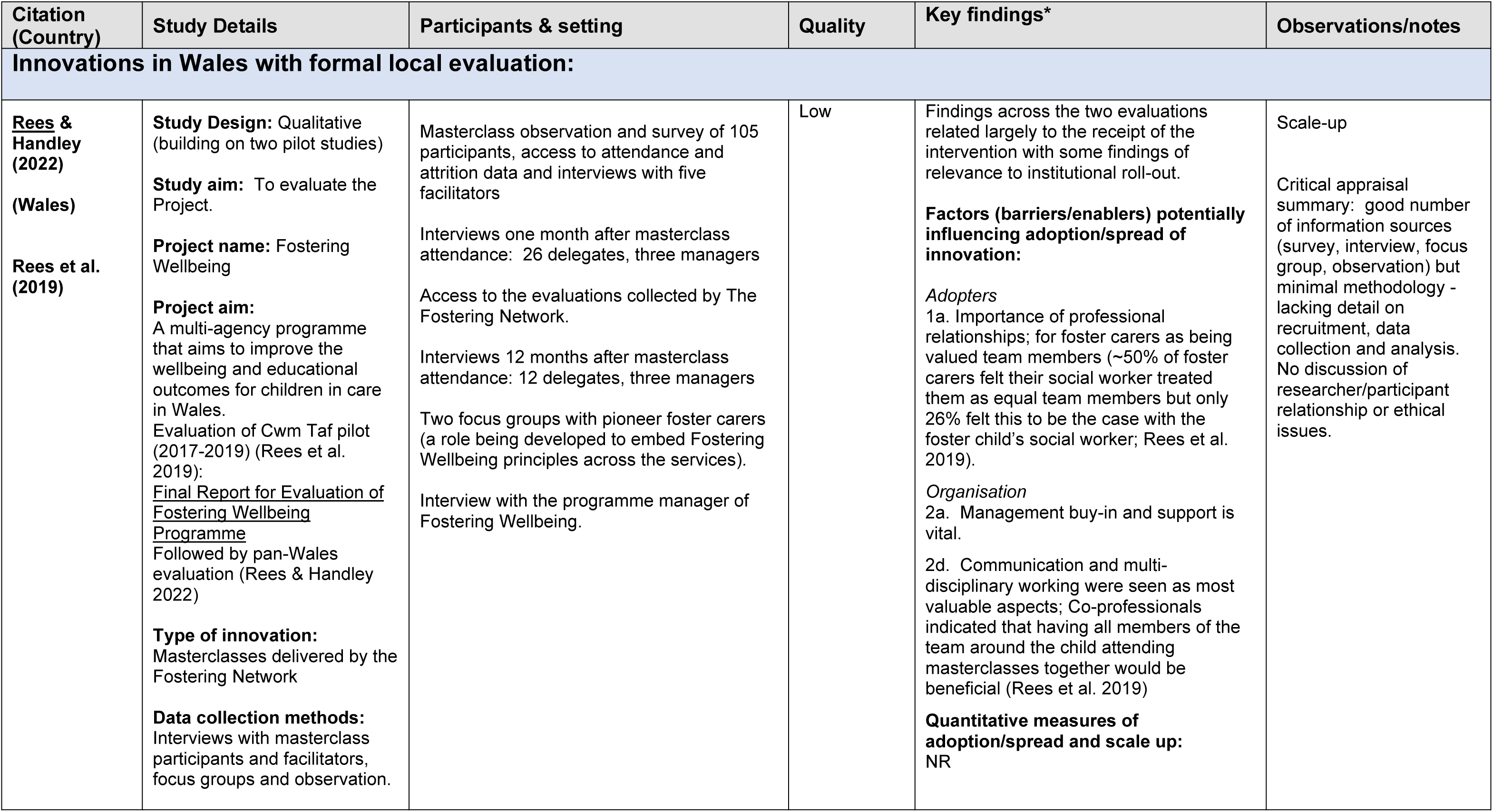
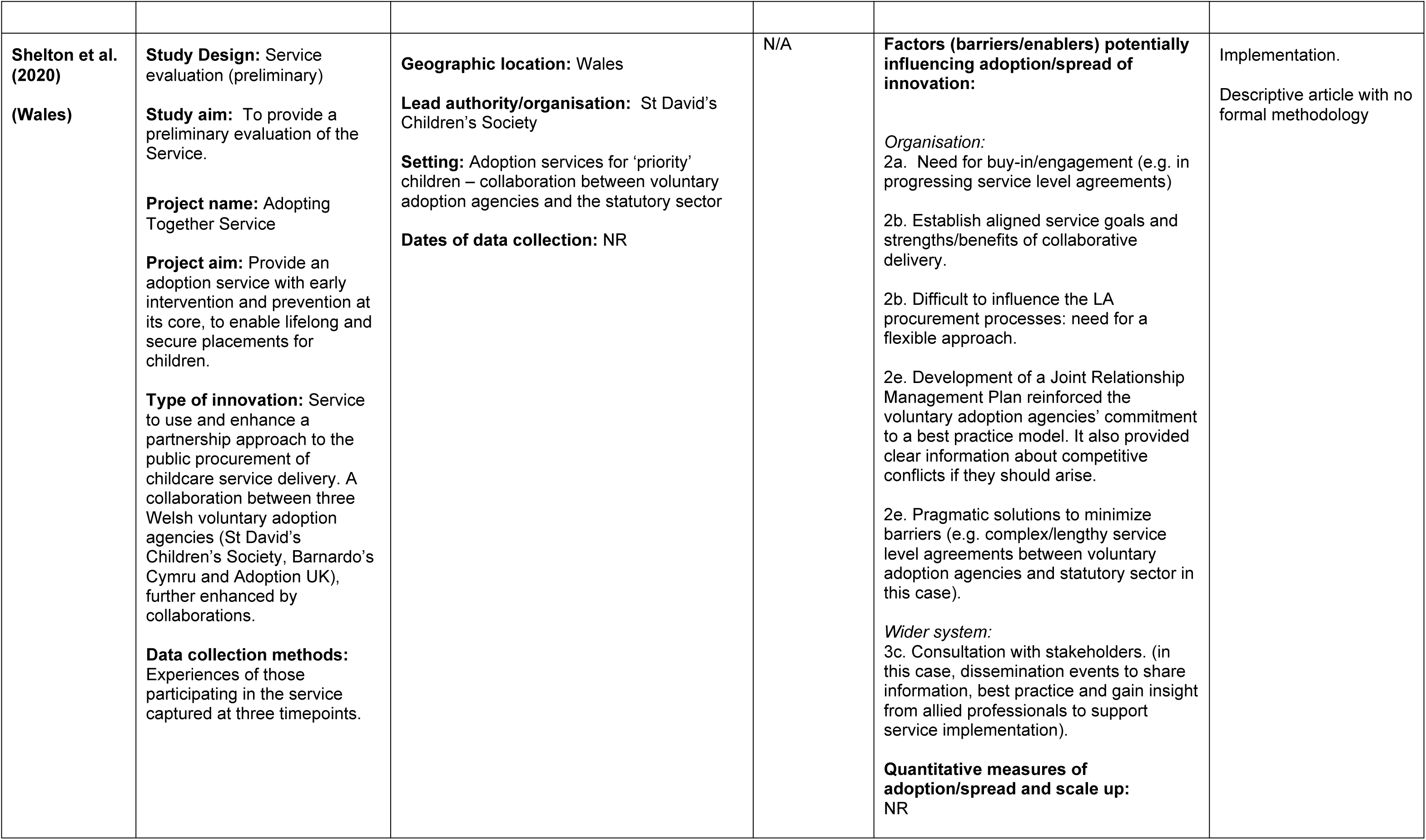
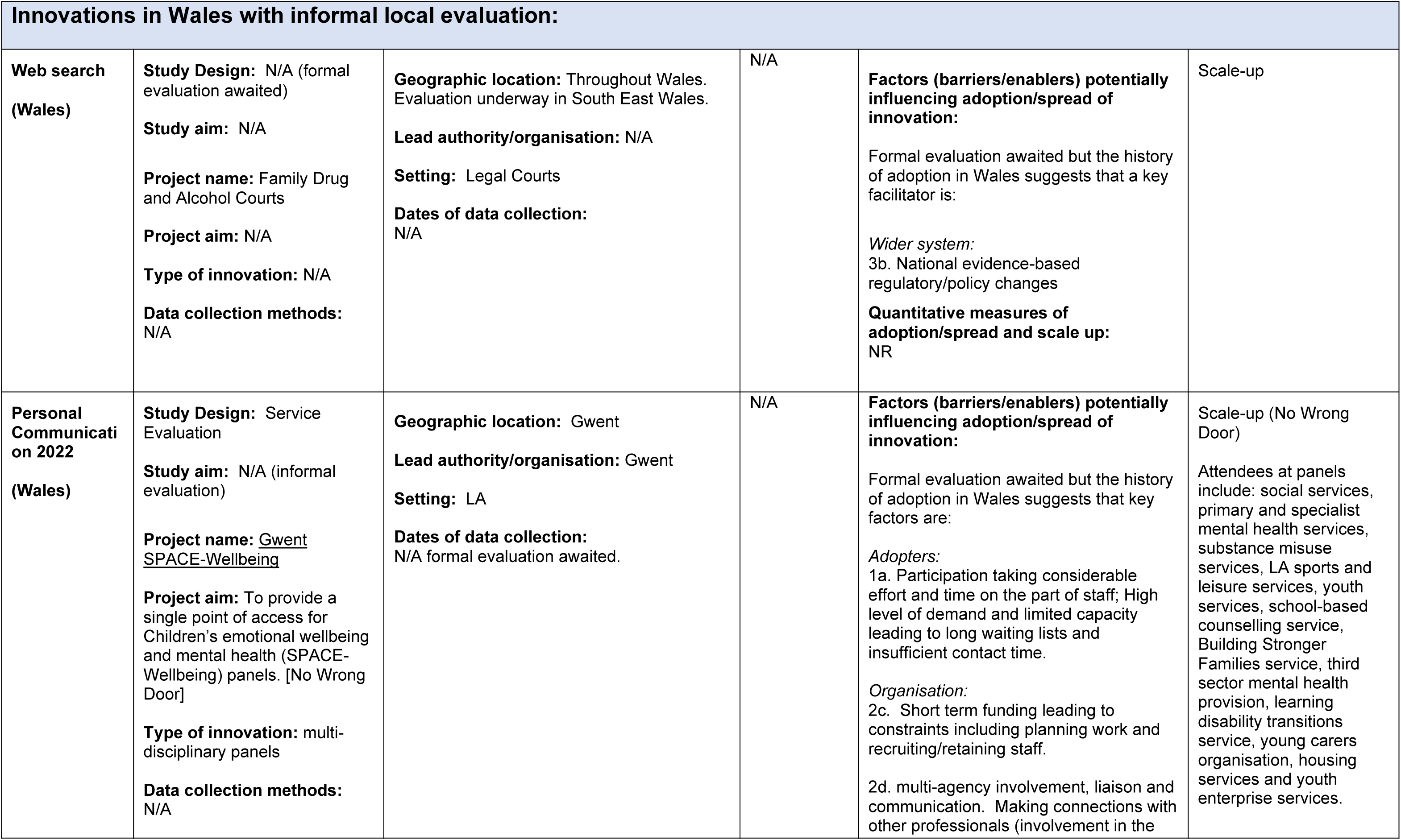
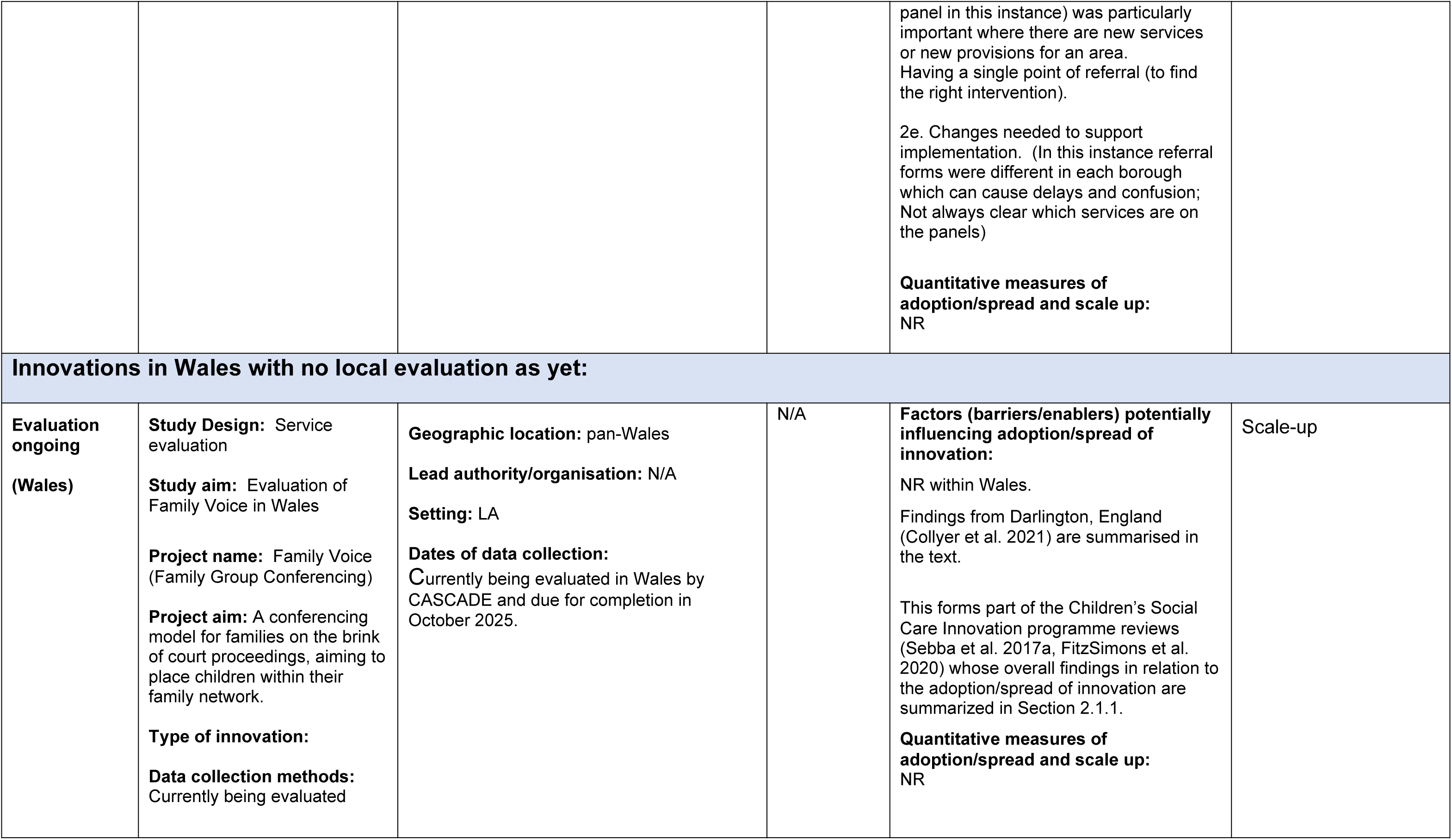
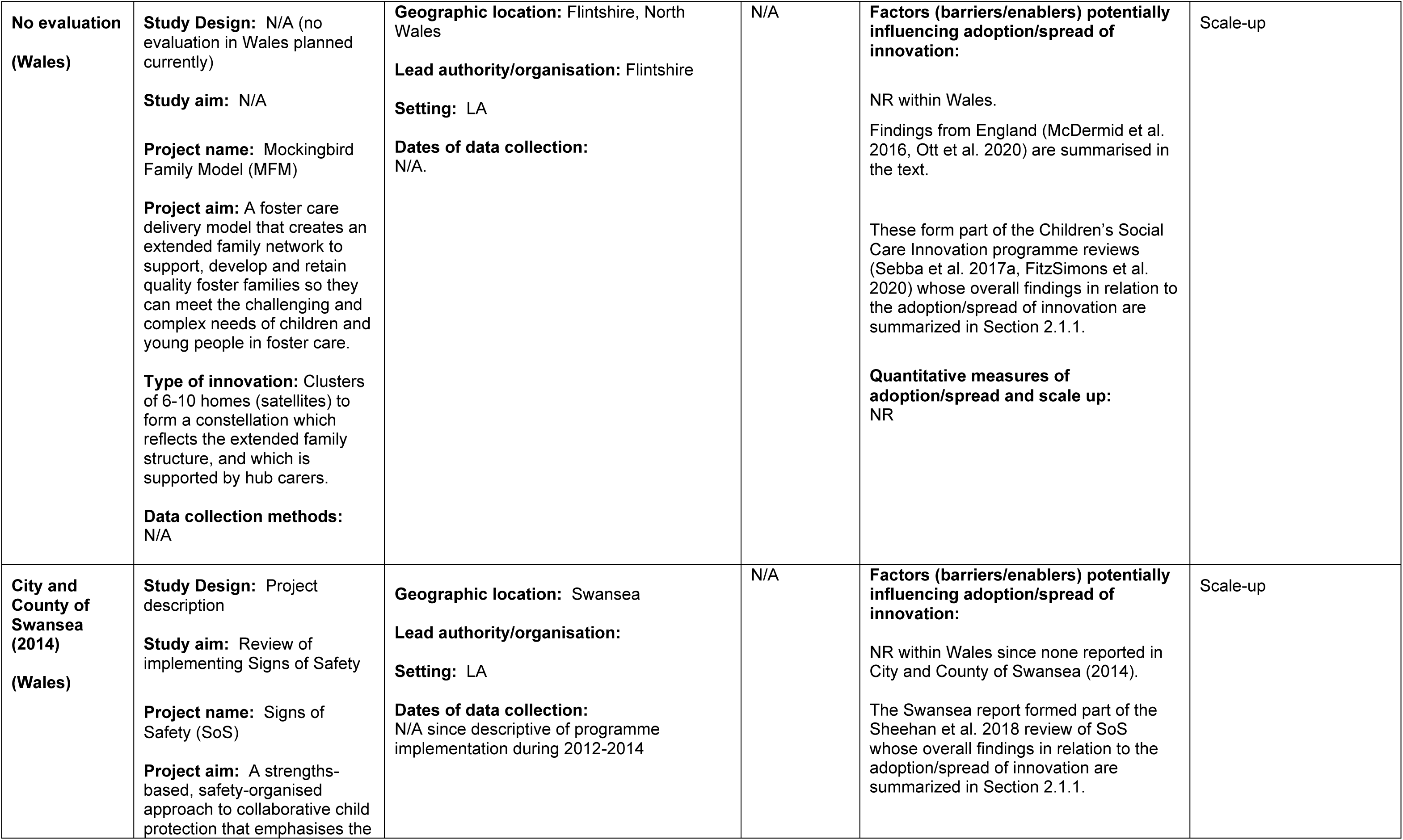
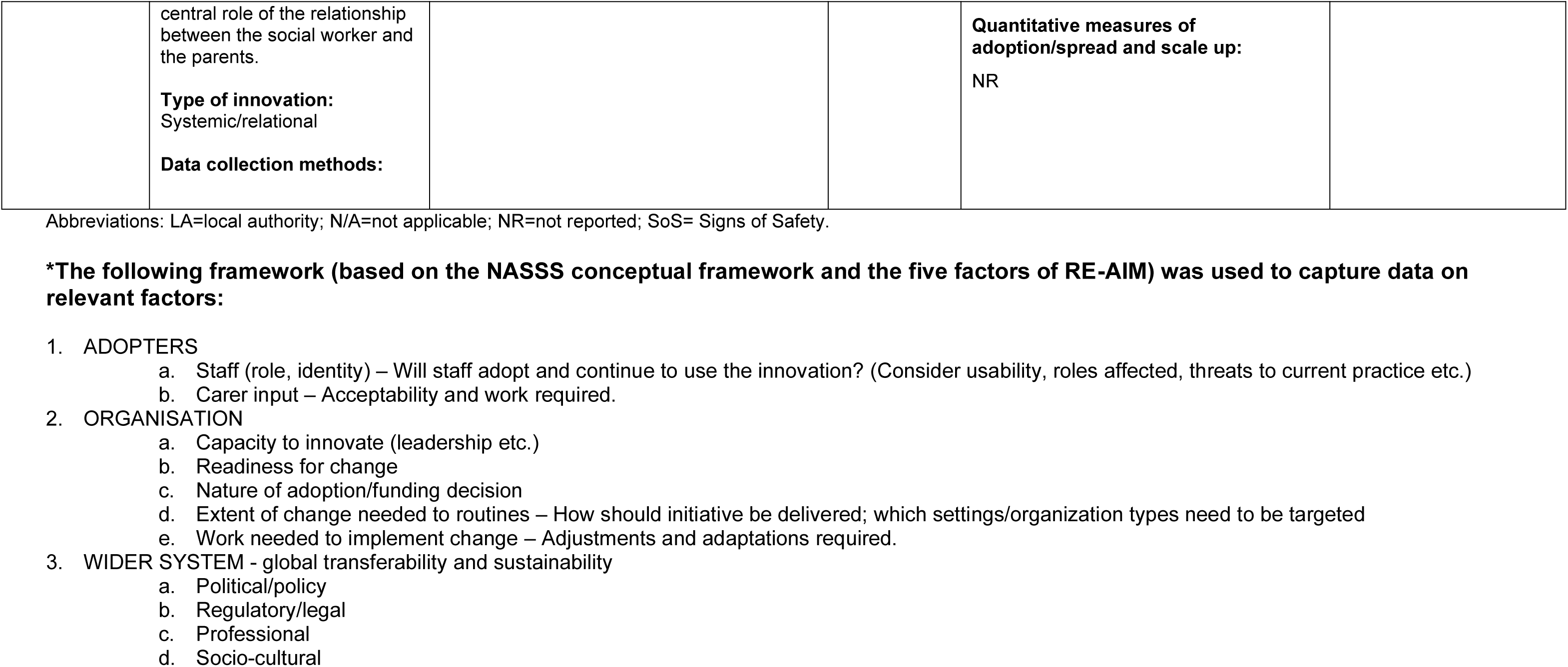
Summary of innovations in Wales.

### 6.3 Quality appraisal tables

See Tables 3-5 in Section 6.2 for a summary of the quality appraisal of each study.

### 6.4 Information available on request

The protocol, all search strategies, details of excluded studies and individual critical appraisal forms (for the five studies that could be formally appraised) are available from

## 7. ADDITIONAL INFORMATION

### 7.1 Conflicts of interest

The authors declare they have no conflicts of interest to report.

## 7.2 Acknowledgements

The SURE team would like to thank stakeholders for their involvement, time and expertise in this rapid review process: Lisa Trigg, Stephanie Griffith, Emma Taylor-Collins, Sara Evans, Rhiannon Evans, Nina Maxwell, Rashmi Kumar.

## 7.3 Disclaimer

The views expressed in this publication are those of the authors, not necessarily Health and Care Research Wales. The Health and Care Research Wales Evidence Centre and authors of this work declare that they have no conflict of interest.

## 8. ABOUT THE HEALTH AND CARE RESEARCH WALES EVIDENCE CENTRE

The Health and Care Research Wales Evidence Centre integrates with worldwide efforts to synthesise and mobilise knowledge from research.

We operate with a core team as part of [Health and Care Research Wales](https://healthandcareresearchwales.org/about-research-community/wales-covid-19-evidence-centre), Welsh Government, and are led by [Professor Adrian Edwards of Cardiff University](https://www.cardiff.ac.uk/people/view/123022-edwards-adrian).

The core team of the centre works closely with collaborating partners in the [Bangor Institute for Health and Medical Research (BIHMR)](https://www.bangor.ac.uk/medical-health-sciences/research), Bangor University, which includes the [Centre for Health Economics and Medicines Evaluation (CHEME)](https://cheme.bangor.ac.uk/) working in collaboration with Health and Care Economics Cymru, [Health Technology Wales](https://healthtechnology.wales/), [Public Health Wales Evidence Service](https://phw.nhs.wales/services-and-teams/observatory/evidence/), [Population Data Science, Swansea University](https://popdatasci.swan.ac.uk/about-us/) using [SAIL Databank](https://saildatabank.com/), the [Wales Centre for Evidence Based Care (WCEBC)](https://www.cardiff.ac.uk/healthcare-sciences/about-us/engagement/the-wales-center-for-evidence-based-care-wcebc), the [Specialist Unit for Review Evidence (SURE)](https://www.cardiff.ac.uk/specialist-unit-for-review-evidence) and [CASCADE](https://cascadewales.org/), Cardiff University.

**Director:** Professor Adrian Edwards

**Website:** www.researchwalesevidencecentre.co.uk

## 9. APPENDICES

## Appendix 1: Resources searched

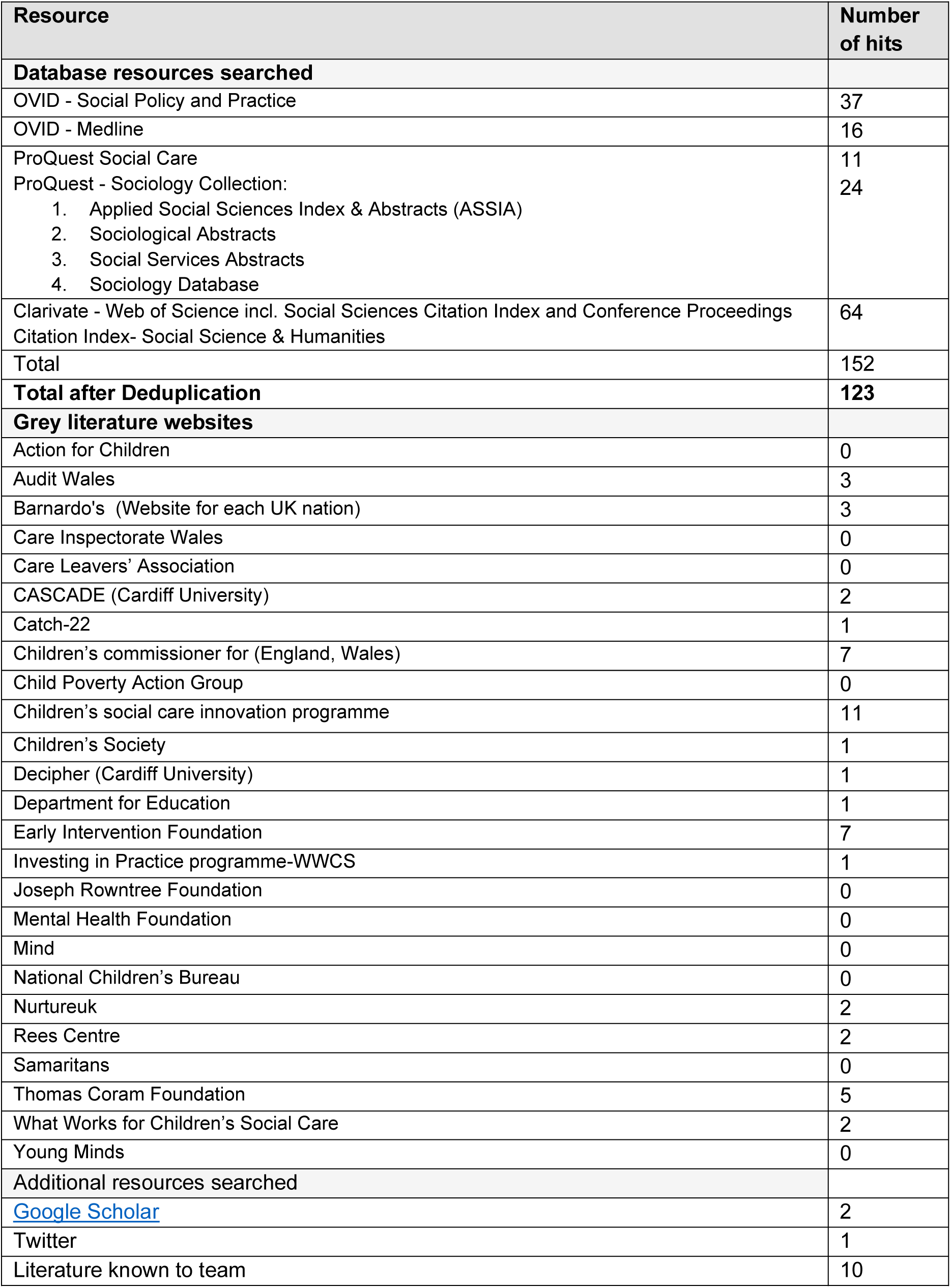

## Appendix 2: Search Strategy

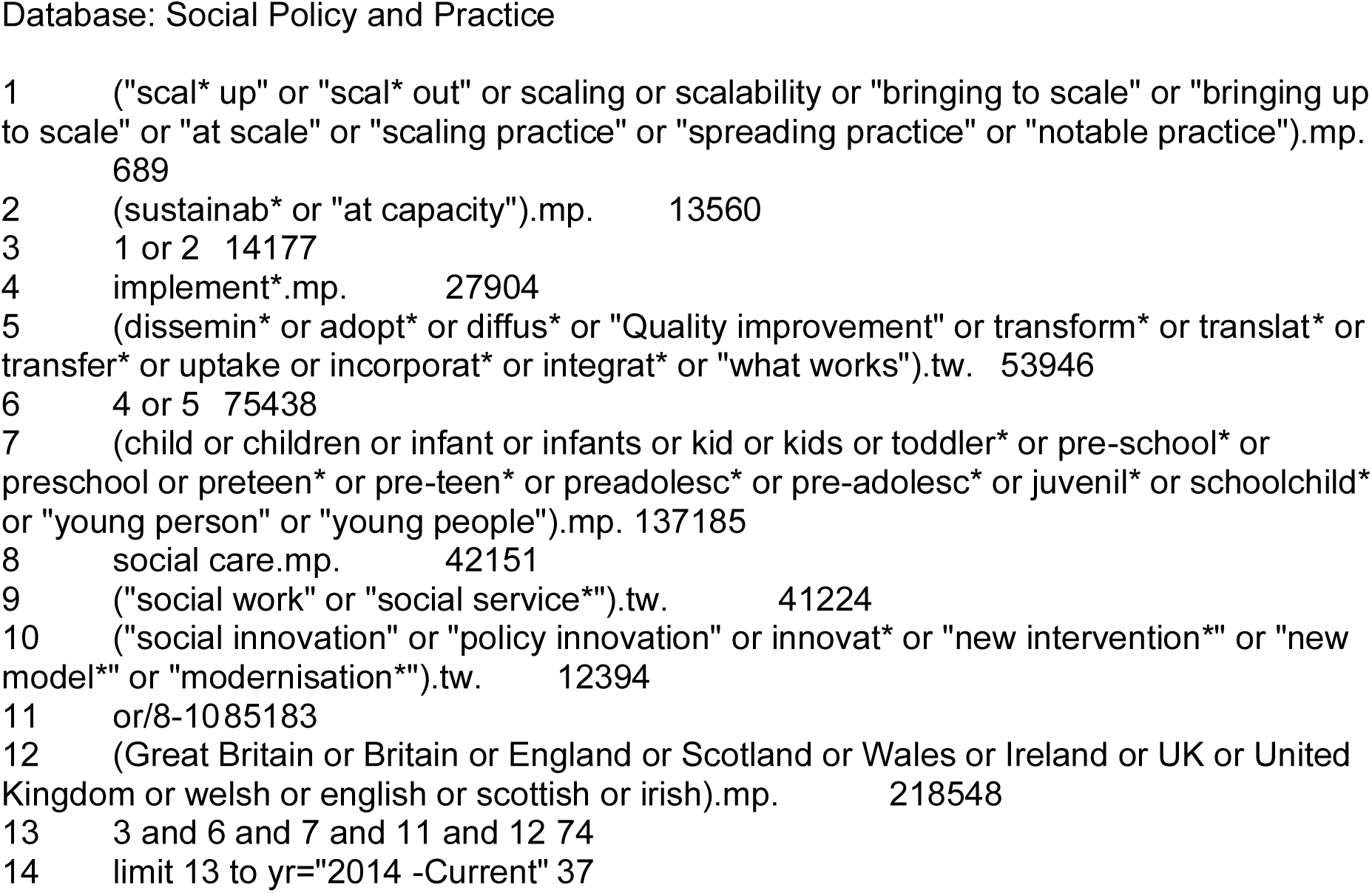

## Abbreviations

CASA: Cognitive and Affective Supervisory Approach
CSCIP: Children’s Social Care Innovation Programme
FDAC: Family Drug and Alcohol Courts
ICF: Integrated Care Fund
LA: Local authority
LSE: London School of Economics
MFM: Mockingbird Family Model
NASSS: Non-adoption, abandonment, scale-up, spread, sustainability framework
NEST: Nurturing, Empowering, Safe, Trusted
RCT: Randomised controlled trial
RE-AIM: Reach, effectiveness, adoption, implementation, maintenance
RPBs: Regional partnership boards
SASCI: Supporting Adult Social Care Innovation Project
SCW: Social Care Wales
SCIE: Social Care Institute for Excellence
SoS: Signs of Safety
PIP: Partners in Practice
VIPP-FC: Positive Parenting and Sensitive Discipline, Foster Care

## Footnotes

1 Sebba J, Luke N, Rees A & McNeish D (2017b). Systemic conditions for innovation in children’s social care. Children’s Social Care Innovation Programme. Thematic Report 4. eISBN: 978-0-9955872-3-6. Available at: https://www.education.ox.ac.uk/wp-content/uploads/2019/06/Systemic-conditions-for-innovation-in-childrens-social-care.pdf

2 Strengths, Weaknesses, Opportunities and Threats.

3 Political, Economic, Social, Technological, Legal and Environmental factors.

4 https://ylab.wales/implementing-mockingbird-family-programme-wales

## Notes

**Funding statement:** The Specialist Unit for Review Evidence was funded for this work by the Health and Care Research Wales Evidence Centre, itself funded by Health and Care Research Wales on behalf of Welsh Government.

### Competing Interest Statement

The authors have declared no competing interest.

### Funding Statement

The Specialist Unit for Review Evidence was funded for this work by the Health and Care Research Wales Evidence Centre, itself funded by Health and Care Research Wales on behalf of Welsh Government

## REFERENCES

Alderson H, Mayrhofer A, Smart D, et al. (2022). An innovative approach to delivering a family-based intervention to address parental alcohol misuse: Qualitative findings from a pilot project. International Journal of Environmental Research and Public Health. 19(13): 8086. doi: https://doi.org/10.3390/ijerph19138086

Callanan S, and Mitchell D. (2020). Scaling innovation in social care. Rapid pragmatic evidence review: summary report. Social Care Institute for Excellence. London, Great Britain. Available at: https://www.scie.org.uk/transformingcare/innovation/network/reports/scaling-innovation [Accessed 30 January 2023].

City and County of Swansea. (2014). Review of implementing Signs of Safety. Solution and safety orientation approach to child protection case work. “Our journey so far”. Swansea: City and County of Swansea, February 2014. https://docplayer.net/6113751-City-and-countyof-swansea-review-of-implementing-signs-of-safety-solution-and-safetyorientation-approach-to-child-protection-case-work.html [Accessed 03 March 2023].

Collyer A, Hennessey A, Sanders M, et al. (2021). Strengthening Families, Protecting Children: Family valued. Pilot Evaluation Report Darlington. London: What Works Centre for Children’s Social Care. Available at: [Accessed 03 March 2023].

Critical Appraisal Skills Programme (2018). CASP (Qualitative) Checklist. Available at: file:///C:/Users/wlbmkm/Downloads/CASP-Qualitative-Checklist-2018_fillable_form%20(1).pdf [Accessed 03 March 2023].

Ecorys UK with Ipsos MORI, Morris K and Family Lives (2017). Dundee Early Intervention Team: independent evaluation report. Available at: https://www.iriss.org.uk/sites/default/files/201711/Dundee%20individual%20report%20FINAL%20REPORT%20260517.pdf [Accessed 03 March 2023].

FitzSimons A, and McCracken K. (2020). Children’s Social Care Innovative Programme Round 2 Final Report. The Department for Education. Available at: https://www.gov.uk/government/publications/childrens-social-care-innovation-programme-final-evaluation-report [Accessed 03 March 2023].

Greenhalgh T, Wherton J, Papoutsi C, et al. (2017). Beyond adoption: A new framework for theorizing and evaluating nonadoption, abandonment, and challenges to the scale-up, spread, and sustainability of health and care technologies. Journal of Medical Internet Research. 19(11): e367. doi: https://doi.org/10.2196/jmir.8775

Godar R, and Botcherby S. (2021). Learning from the Greater Manchester Scale and Spread Programme: Spreading innovation across a city-region Final Overview Report. Research in Practice; Darlington Trust. Available at: https://www.researchinpractice.org.uk/media/5568/scale_and_spread_report_v4.pdf [Accessed 03 March 2023].

McDermid S, Baker C, Lawson, D, et al. (2016). The evaluation of the Mockingbird Family Model. Final evaluation report. Children’s Social Care Innovation programme evaluation, 4. Available at: https://www.gov.uk/government/publications/mockingbird-family-model-evaluation [Accessed 03 March 2023].

National Assembly Wales (2014). Social Services and Well-being (Wales) Act 2014. Available at: https://www.legislation.gov.uk/anaw/2014/4/resources [Accessed 03 March 2023].

Oliveira P, Stevens E, Barge L, et al. (2022). A modified video-feedback intervention for carers of foster children aged 6 years and under with reactive attachment disorder: a feasibility study and pilot RCT. Health Technology Assessment (Winchester, England). 26(35): 1–106. doi: https://doi.org/10.3310/SLIZ1119

Ott E, McGrath-Lone L, Pinto V, et al. (2020). Mockingbird Programme: Evaluation Report. London: Department for Education. Available at: https://assets.publishing.service.gov.uk/government/uploads/system/uploads/attachment_data/file/933119/Fostering_Network_Mockingbird.pdf [Accessed 03 March 2023].

Plumridge G, and Sebba J. (2018). Rees Centre: Evaluation of Birmingham City Council’s1 Step Down Programme Report of the Findings September. https://www.education.ox.ac.uk/wp-content/uploads/2019/05/285171.pdf [Accessed 03 March 2023].

Rees A, Maxwell N, Grey J, et al. (2019). Final Report for Evaluation of Fostering Wellbeing Programme 2019. Available at: https://orca.cardiff.ac.uk/id/eprint/128577/1/Final%20Report%20for%20Evaluation%20of%20Fostering%20Wellbeing%20Programme%205th%20December.pdf [Accessed 03 March 2023].

Rees A, and Handley B. (2022). Evaluation of fostering wellbeing. Available at: https://orca.cardiff.ac.uk/id/eprint/154327/1/Final%20version%2014th%20September15%20%281%29.pdf [Accessed 03 March 2023].

Ruch G, and Maglajlic RA. (2020). Partners in Practice: practice review report. Available at: https://assets.publishing.service.gov.uk/government/uploads/system/uploads/attachment_data/file/932330/Partners_in_Practice_PiP_Features.pdf [Accessed 03 March 2023].

Shea BJ, Reeves BC, Wells G, et al. (2017). AMSTAR 2: a critical appraisal tool for systematic reviews that include randomised or non-randomised studies of healthcare interventions, or both. BMJ (Clinical research ed.). 358: j4008. doi: https://doi.org/10.1136/bmj.j4008

Sheehan L, O’Donnell C, Brand SL, et al. (2018). Signs of Safety: Findings from a mixed-methods systematic review focussed on reducing the need for children to be in care. London: What Works Centre for Children’s Social Care. Available at: https://whatworks-csc.org.uk/wp-content/uploads/Signs_of_Safety_a_mixed_methods_systematic_review.pdf [Accessed 03 March 2023].

Sebba J, Luke N, McNeish D, et al. (2017a). Children’s Social Care Innovation Programme. Final evaluation report. Available at: https://assets.publishing.service.gov.uk/government/uploads/system/uploads/attachment_data/file/659110/Children_s_Social_Care_Innovation_Programme_-_Final_evaluation_report.pdf [Accessed 03 March 2023].

Sebba J, Luke N, Rees A, et al. (2017b). Systemic conditions for innovation in children’s social care: Children’s Social Care Innovation Programme, thematic report 4. Available at: www.education.ox.ac.uk/wp-content/uploads/2019/06/Systemic-conditions-for-innovation-in-childrens-social-care.pdf [Accessed 03 March 2023].

Shaw RB, Sweet SN, McBride CB, et al. (2019). Operationalizing the reach, effectiveness, adoption, implementation, maintenance (RE-AIM) framework to evaluate the collective impact of autonomous community programs that promote health and well-being. BMC Public Health. 19(1): 803. doi: https://doi.org/10.1186/s12889-019-7131-4

Shelton KH, Merchant C, Lynch J. (2020). The Adopting Together Service: how innovative collaboration is meeting the needs of children in Wales waiting the longest to find a family. Adoption & Fostering. 44(2): 128–141. doi: https://doi.org/10.1177/0308575920920390

Turney D and Ruch G. (2018). What makes it so hard to look and to listen? Exploring the use of the Cognitive and Affective Supervisory Approach with children’s social work managers. Journal of Social Work Practice. 32:2: 125–138. doi: 10.1080/02650533.2018.1439460

Zhang S, Huang H, Wu Q, et al. (2019). The impacts of family treatment drug court on child welfare core outcomes: A meta-analysis. Child Abuse & Neglect. 88: 1–14. doi: 10.1016/j.chiabu.2018.10.014

Zigante V, Malley J, Boaz A, et al. (2022). How can the adult social care sector develop, scale and spread innovations? A review of the literature from an organisational perspective. Care Policy and Evaluation Centre; The London School of Economics and Political Science. Available at: https://www.sasciproject.uk/publications [Accessed 03 March 2023].

